# Accuracy of screening tests for cervical pre-cancer in women living with HIV in low-resource settings: a paired prospective study in Lusaka, Zambia

**DOI:** 10.1101/2023.05.31.23290779

**Authors:** Katayoun Taghavi, Misinzo Moono, Mulindi Mwanahamuntu, Marie Roumet, Andreas Limacher, Herbert Kapesa, Thamsanqa Madliwa, Anne Rutjes, Partha Basu, Nicola Low, Albert Manasyan, Julia Bohlius

## Abstract

**Introduction:** To provide evidence to improve cervical screening for women living with HIV (WLHIV), we assessed the accuracy of screening tests that can be used in low-resource settings and give results at the same visit.

**Methods:** We conducted a paired, prospective study among consecutive eligible WLHIV, aged 18–65 years, receiving cervical cancer screening at one hospital in Lusaka, Zambia. The histopathological reference standard was multiple biopsies taken at two time points. The target condition was high-grade cervical intraepithelial neoplasia (CIN2+). The index tests were high-risk human papillomavirus detection (hrHPV, Xpert HPV, Cepheid), portable colposcopy (Gynocular, Gynius), and visual inspection with acetic acid (VIA). Accuracy of stand-alone and test combinations were calculated as the point estimate with 95% confidence intervals. A sensitivity analysis considered disease when only visible lesions were biopsied.

**Results:** Among 371 participants with histopathological results, 27% (101/371) women had CIN2+ and 23% (23/101) was not detected by any index test. Sensitivity and specificity for stand-alone tests were: hrHPV, 67.3% (95% CI: 57.7–75.7) and 65.3% (59.4–70.7); Gynocular 51.5% (41.9–61.0) and 80.0% (74.8–84.3); and VIA 22.8% (15.7–31.9) and 92.6% (88.8–95.2), respectively. The combination of hrHPV testing followed by Gynocular had the best balance of sensitivity (42.6% [33.4–52.3]) and specificity (89.6% [85.3–92.7]). All test accuracies improved in sensitivity analysis.

**Conclusion:** The low accuracy of screening tests assessed might be explained by our reference standard, which reduced verification and misclassification biases. Better screening strategies for WLHIV in low-resource settings are urgently needed.

**Registration number:** The trial was registered prospectively at ClinicalTrials.gov (ref: NCT03931083). The study protocol has been previously published, and the statistical analysis plan can be accessed on ClinicalTrials.gov.

**Key messages:** *What is already known on this topic:* The 2021 World Health Organization guidelines recommend that women living with HIV (WLHIV) receive screening for high risk human papillomavirus high-risk human papillomavirus (hrHPV) genotypes at three- to five-year intervals, followed by a triage test to determine whether treatment is needed but this is based on low and moderate certainty evidence.

*What this study adds:* This study among WLHIV in Lusaka, Zambia evaluated three screening tests that allow same-day treatment; hrHPV test, portable colposcopy (Gynocular), and visual inspection with acetic acid (VIA), using strict methods to reduce verification and misclassification biases. The test accuracy of the different screening was poor, with sensitivities and specificity for stand-alone tests: hrHPV, 67.3% and 65.3%; Gynocular 51.5% and 80.0%; and VIA 22.8% and 92.6%; respectively.

*How this study might affect research, practice or policy:* Our findings have implications for research and cervical cancer screening policies among WLHIV if test-accuracy in this high-risk population has been overestimated from a majority of exsisting studies that are affected by verification and misclassification biases. Methodologically robust studies are crucial to inform cervical cancer screening practices and policies for the successful implementation of a cervical cancer elimination plan in sub-Saharan Africa, where 85% of women with cervical cancer and HIV live.

## Introduction

The World Health Organization (WHO) strategy to eliminate cervical cancer aims to improve prevention and treatment among women living with HIV infection (WLHIV).^1^ A conditional recommendation for WLHIV suggests testing for high-risk human papillomavirus (hrHPV) followed by an additional screening test based on moderate certainty evidence.^1^ Cervical cancer remains the leading cause of cancer-related death among women in sub-Saharan African (SSA) countries, where more than half of cervical cancer cases are attributable to HIV.^2^ Increased life expectancy on antiretroviral therapy (ART) increases the number of women with persistent hrHPV infection, which may progress to cervical pre-cancer and cancer.^3-5^

In low-resource settings, tests that give same-day results and lead to decisions about treatment are preferred. An evaluation of alternative same-day screening tests among WLHIV, using methods that minimise verification biases, has not yet been conducted.^1,6,7^ Colposcopy is the cornerstone of visual assessment for cervical cancer screening, used in screening pathways of high-resource countries but is rarely accessible in low-resource settings where most WLHIV live. In low-resource settings, visual inspection with acetic acid (VIA) is commonly used,^5,6^ but with low accuracy, particularly for WLHIV.^7,8^ The WHO strategy recommends molecular tests to detect hrHPV, which were reported to have a sensitivity of 91.6% (95% CI 88.1–94.1) among WLHIV in a systematic review.^7^ Our objectives were to assess the accuracy of molecular and visual screening tests (hrHPV testing, portable colposcopy using the Gynocular, and VIA).

## Methods

This study followed a published protocol^9^ and is reported according to the STARD 2015 guideline (Appendix, S1). Ethical approval was granted by the National Health Research Authority and the University of Zambia Biomedical Research Ethics Committee (ref: 014-09-18), the Zambia Medicines Regulatory Authority (ref: DMS/7/9/22/CT/084), the International Agency for Research on Cancer (IEC project number 18-15), and Swissethics (ref: 2018-01399).

### Study design and participants

We conducted a single-site, paired (all women received all tests) prospective test-accuracy study among WLHIV in Lusaka, Zambia. We assessed women presenting consecutively to the cervical cancer screening clinic at Kanyama General Hospital for eligibility. We enrolled women aged 18 to 65 years with confirmed HIV infection who had ever had sex, gave written consent, and agreed to return for a six-month follow-up visit. We excluded women with a history of cervical cancer or total hysterectomy and those vaccinated against HPV. Women enrolled were a consecutive series who fulfilled eligibility criteria and for whom the research staff could complete all study procedures.

### Procedures

Two nurses and one research assistant collected and tested specimens. They performed procedures in separate rooms and documented findings on separate case-report forms. Clinical team members did not discuss results, and participants were asked not to communicate findings to staff. After consent, a nurse recorded medical history and sociodemographic information. Blood tests for HIV RNA viral load (Cobas HIV-1/2 Qual; Roche Molecular Systems, New Jersey, USA) and CD4 cell count (Pima CD4 Analyzer, Alere, Waltham, USA) were taken at baseline.

### Reference standard

The target condition was the histological presence of cervical intraepithelial neoplasia grade two and above (CIN2+) or high-grade squamous intraepithelial lesions (HSIL) at baseline or six-month follow-up. A study nurse took biopsies during the colposcopy examination. If lesions were seen, she took at least two biopsies from those that looked the most severe. If no lesions were seen, she took four biopsies from clock-face positions 3, 6, 9, and 12 o’clock within the transformation zone. The nurse received training in colposcopy and biopsy-taking from a gynaecologist based at the International Agency for Research on Cancer (IARC) and a local senior gynaecologist (S2). To further reduce detection bias in the reference standard, we took a second set of biopsies from each woman six months later to identify cases of disease missed at baseline. Biopsies were assessed histologically at two independent laboratories in South Africa and Zambia. An expert gynaecological pathologist in each laboratory, blinded to the clinical findings, examined all biopsies and classified them using the Bethesda squamous intraepithelial lesion system.^10^ They reviewed all histopathology results via teleconference and reached an agreement on diagnosis. Any sample with CIN2 or ambiguous findings was tested with p16 immunostaining.^10,11^ We dichotomised histopathological findings into low-grade and HSIL by the lower anogenital squamous terminology definitions and the WHO Classification of Tumours of Female Reproductive Organs.^10^

All women with VIA-positive findings, or CIN2+ or HSIL on histology, were offered treatment with cryotherapy, thermoablation, or loop electrosurgical excision procedure (LEEP) as clinically indicated. Women with histopathologically confirmed cervical cancer were referred to the University Teaching Hospital in Lusaka for treatment.

### Index tests

A trained nurse (<u>S2</u>) did a speculum examination and collected specimens, followed by VIA. An endocervical sample was taken using a single-use cytobroom and immediately placed into ThinPrep PreservCyt solution (Hologic, Marlborough, Massachusetts, USA) (S2). A swab from the posterior vaginal fornix was also taken and tested for *T. vaginalis*. A research assistant processed both specimens within two to four hours of collection using the GeneXpert platform (Cepheid, Sunnyvale, California, USA) at the study site, as per the manufacturers’ instructions. The Xpert HPV test detects 14 hrHPV subtypes, categorised for reporting as HPV16, HPV18/45 (subtypes 18 and/or 45), and HPV other (any of subtypes 31, 33, 35, 39, 51, 52, 56, 58, 59, 66, and 68).

VIA examination followed IARC methodology (S2).^12^ As per local guidelines, VIA nurses categorised indeterminate findings as abnormal. In a separate room, a different nurse performed a colposcopic examination using the Gynocular (S3) following methods described in the IARC colposcopy manual.^14^ We used the Swede score to standardise the documentation of findings on visual inspection with a score from 0 (abnormality not seen) to 2 (most severe)^13^ (S4).

### Interpretations of results

The histological presence of CIN2+ or HSIL at baseline or six-month follow-up was considered as the disease outcome in the primary analysis. New cases of CIN2+ at follow-up were considered as diagnoses that had been missed at baseline. A positive hrHPV test result was defined as the detection of any of the 14 subtypes detected by the Xpert HPV test. VIA findings were dichotomised as positive (abnormal, suspicious of cancer, or indeterminate) and negative (normal). We used receiver operating characteristic (ROC) curve analysis to calculate the area under the curve (AUC) for each level of the Swede score as assessed by Gynocular colposcopy. We then used the Youden cut-off in the primary analysis, optimising both sensitivity and specificity. In an additional analysis, we used cut-offs maximising either sensitivity (≥90%) or specificity (≥90%).

### Sample size and statistical analyses

We required a sample of 350 participants based on estimates of precision for the sensitivity and specificity of Gynocular, hrHPV, and VIA as stand-alone tests for detecting CIN2+ lesions (S5). We aimed to recruit 450 women to allow for incomplete data.

In our analyses, we used consensus agreement of the reference standard. We assessed agreement between the two pathologists for the reference standard using Cohen’s kappa coefficient (κ). The accuracy measures used to assess Gynocular, VIA, and hrHPV tests were sensitivity and specificity, positive and negative predictive values, positive and negative likelihood ratios, false positive and false negative rates, and diagnostic odds ratios (DORs). Screening test accuracy measures were estimated with 95% Wilson CIs. Using the same approach, we evaluated the accuracies of two tests used together. We considered the combination positive if both single tests were positive, and negative otherwise. This mimics the clinical scenario where the second test is used to decide whether treatment is required (triage test).^1^

We also described test accuracy measures in subgroups defined by age (<25, 26–35, 36–45, >46 years, and menopausal status), parity, ART status, co-infection with *T. vaginalis*, methods of contraception, and CD4 cell count. Sensitivity and specificity were calculated for each subgroup. Estimated sensitivity values were compared with those found in the reference category by calculating the sensitivity ratio. To investigate the occurrence of effect modification by patient characteristics on the association between the diagnostic test and disease status, we used univariable and multivariable logistic regression models and tested for the interaction between the diagnostic test and patient characteristics on disease status. We considered the following patient’s characteristics: age, menopause, parity, ART status, *T. vaginalis* at baseline, methods of contraception, HIV RNA, CD4 cell count, history of treatment for pre-cancer, and education level. Adjustment was performed considering all before mentioned patient characteristics as predictors and performing a stepwise model selection based on the Akaike information criterion (AIC).

We conducted the following sensitivity analyses to investigate the influence of unverifiable assumptions. First, we explored a possible training effect by assessing the first 10% of participants separately. Second, we explored the impact of the COVID-19 pandemic by conducting primary analyses separately on women who finished the study before 28 March 2020 (study ceased due to the pandemic). Third, we assessed the impact of missing or indeterminate results in the reference standard or screening test results by considering them first as positive cases, then as negative. Finally, acknowledging that biopsy and HPV tests may be performed and interpreted differently, we conducted analyses using a reference standard from a hypothetical scenario in which a biopsy was taken only from visible lesions and using different categories of hrHPV test results.

## Results

### Flow of participants

Between May 2019 and March 2021, we assessed 413 women, enrolled 376, and included 375 in the analysis (one woman was found to have had a total hysterectomy; Figure 1). We had valid reference standard results for 371 women. VIA and hrHPV tests were performed on the 375 enrolled with Gynocular examination conducted on 373. Before the COVID-19 pandemic stopped the study, 104 completed the follow-up. The follow-up period for deriving the reference standard was six months. There were no adverse events.

**Figure 1:**
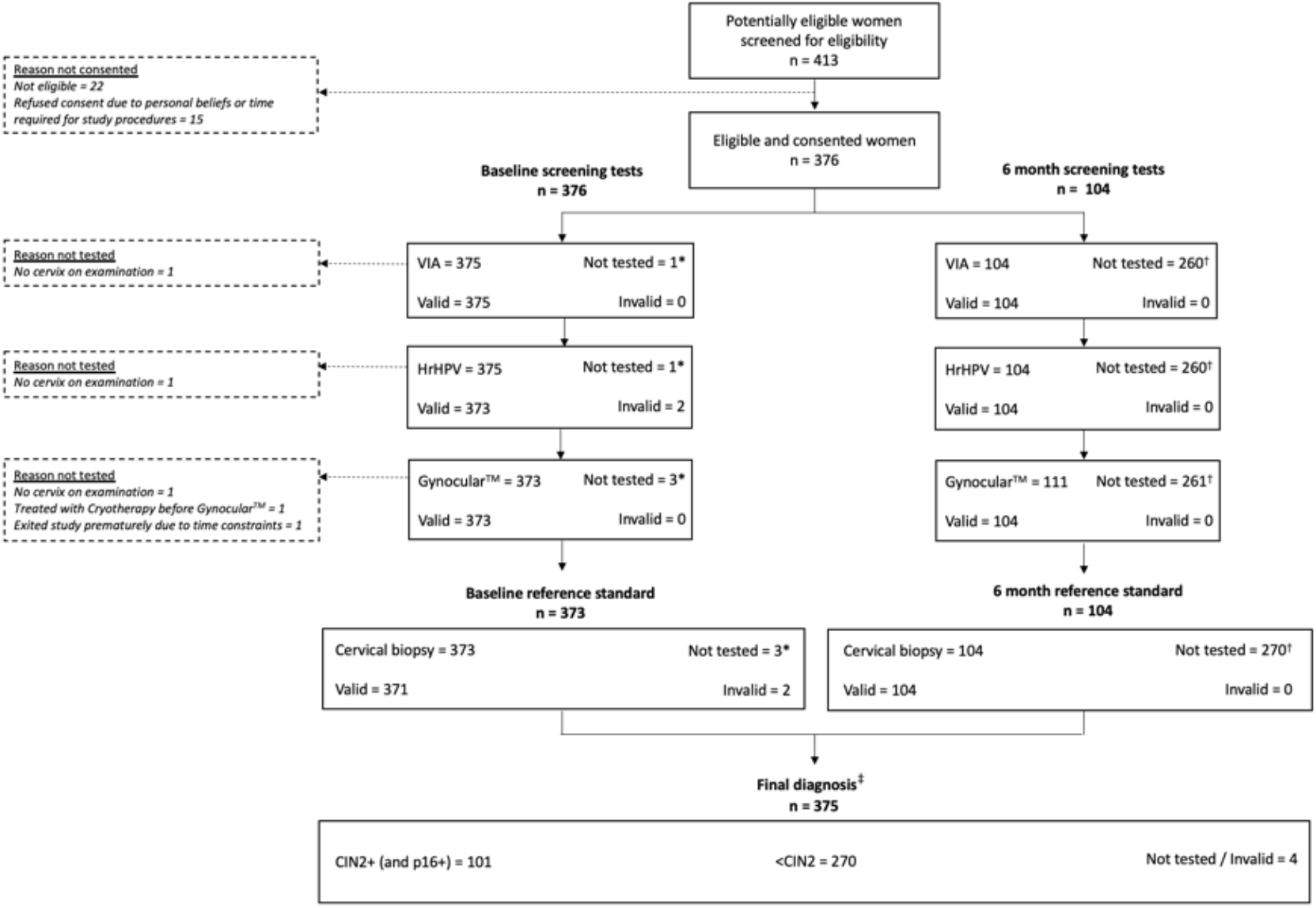
Flow of participants. Diagram to show the number of women receiving screening tests and reference standard, and analysed in the study. Data are n= number of women. VIA= visual inspection of the cervix with acetic acid. HrHPV= high-risk human papillomavirus. CIN2+= cervical intraepithelial neoplasia grade two and above. p16+= expression of cell cycle regulatory protein 16^INK4A^. <CIN2= cervical intraepithelial neoplasia grade one and below. *= excluded from the analysis as she did not receive any of the study screening tests. †= did not receive further tests as the study stopped following the COVID-19 pandemic. ‡= the final diagnosis used in the analyses considers histopathological diagnosis for all women at baseline or six-month follow-up, disease (CIN2+) was considered as present when biopsies from at least one time-point were positive.

### Patient characteristics

The full baseline characteristics are in Table 1. At enrolment, participants had a median age of 37 years (interquartile range 31–44) and median parity of three (IQR 2–5). Most were not using any contraception (62%, n=231), did not smoke (99%, n=373) or use insunko^16^ (a smokeless, carcinogenic tobacco product that can be used vaginally; 95%, n=355), and did not drink alcohol (88%, n=331). Most had never undergone cervical cancer screening (71%, n=267); VIA was the modality among those who had received screening. Seven women (2%) had received previous cryotherapy treatment. Almost all were on ART (99%, n=374) and had well-controlled HIV infection. Women with histological CIN2+ were more likely to have a CD4 cell count <200 per mm^3^ (7/101, 7%) than women without (5/270, 2%) and viral load ≥50 copies/mL (22/101, 22% and 36/270, 13%, respectively). We report baseline characteristics from routinely available data of all women aged 18 to 65 years seen at Kanyama HIV clinic in S6.

**Table 1:** Baseline characteristics.

| Patient characteristics | Overall<br>n = 375 | CIN2+<br>n = 101 | <CIN2<br>n = 270 | Not tested/Invalid<br>n = 4 |
| --- | --- | --- | --- | --- |
| Age |  |  |  |  |
| Median (*IQR*) | 37 (31, 44) | 36 (28, 42) | 37 (31, 44) | 46 (42, 54) |
| Missing | 7 | 2 | 5 | 0 |
| Menopause |  |  |  |  |
| No | 321 (86%) | 90 (89%) | 229 (85%) | 2 (50%) |
| Yes | 54 (14%) | 11 (11%) | 41 (15%) | 2 (50%) |
| Marital status |  |  |  |  |
| In a relationship/Married | 244 (65%) | 70 (69%) | 173 (64%) | 1 (25%) |
| Separated/Divorced | 69 (18%) | 12 (12%) | 55 (20%) | 2 (50%) |
| Single | 12 (3%) | 4 (4%) | 8 (3%) | 0 (0%) |
| Widowed | 50 (13%) | 15 (15%) | 34 (13%) | 1 (25%) |
| Employment |  |  |  |  |
| Working | 157 (42%) | 39 (39%) | 116 (43%) | 2 (50%) |
| Not working | 218 (58%) | 62 (61%) | 154 (57%) | 2 (50%) |
| Education |  |  |  |  |
| Did not finish secondary | 308 (82%) | 86 (85%) | 219 (81%) | 3 (75%) |
| Finished secondary | 54 (14%) | 12 (12%) | 41 (15%) | 1 (25%) |
| More than secondary | 13 (4%) | 3 (3%) | 10 (4%) | 0 (0%) |
| Income* (Kwacha/month) |  |  |  |  |
| None | 216 (58%) | 61 (60%) | 153 (57%) | 2 (50%) |
| 1–500 | 50 (13%) | 16 (16%) | 34 (13%) | 0 (0%) |
| 501–1000 | 74 (20%) | 15 (15%) | 57 (21%) | 2 (50%) |
| 1001–2500 | 32 (9%) | 9 (9%) | 23 (9%) | 0 (0%) |
| 2501–5000 | 3 (1%) | 0 (0%) | 3 (1.1%) | 0 (0%) |
| Smoking |  |  |  |  |
| No | 373 (99%) | 101 (100%) | 268 (99%) | 4 (100%) |
| Yes | 2 (1%) | 0 (0%) | 2 (1%) | 0 (0%) |
| Alcohol |  |  |  |  |
| No | 331 (88%) | 92 (91%) | 235 (87%) | 4 (100%) |
| Yes | 44 (12%) | 9 (8.9%) | 35 (13%) | 0 (0%) |
| Insunko |  |  |  |  |
| No | 355 (95%) | 95 (94%) | 256 (95%) | 4 (100%) |
| Yes | 20 (5%) | 6 (6%) | 14 (5%) | 0 (0%) |
| Age at sexual debut |  |  |  |  |
| Median (*IQR*) | 17(16, 19) | 17(15, 19) | 17(16, 19) | 17 (16, 18) |
| Missing | 3 | 0 | 3 | 0 |
| Contraception use |  |  |  |  |
| No | 231 (62%) | 66 (65%) | 161 (60%) | 4 (100%) |
| Yes | 144 (38%) | 35 (35%) | 109 (40%) | 0 (0%) |
| Gravidity |  |  |  |  |
| Median (*IQR*) | 4 (2, 5) | 3 (2, 5) | 4 (2, 5) | 3 (2, 6) |
| Parity |  |  |  |  |
| Median (*IQR*) | 3 (2, 5) | 3 (2, 4) | 4 (2, 5) | 3 (2, 6) |
| On ART |  |  |  |  |
| No | 1 (0.3%) | 0 (0%) | 1 (0.4%) | 0 (0%) |
| Yes | 374 (100%) | 101 (100%) | 269 (100%) | 4 (100%) |
| CD4 cell count (cells/mm3) |  |  |  |  |
| Median (IQR) | 542 (418, 759) | 532 (406, 742) | 543 (421, 763) | 1111 (928, 1123) |
| Missing | 2 | 0 | 1 | 1 |
| HIV RNA load (copies/mL) |  |  |  |  |
| <50 | 316 (84%) | 79 (78%) | 234 (87%) | 3 (100%) |
| 50–1000 | 29 (8%) | 10 (10%) | 19 (7%) | 0 (0%) |
| 1001–10000 | 11 (3%) | 6 (6%) | 5 (2%) | 0 (0%) |
| 10001 and more | 18 (5%) | 6 (6%) | 12 (4%) | 0 (0%) |
| Missing | 1 | 0 | 0 | 1 |
| History of previous cervical screening |  |  |  |  |
| No | 267 (71%) | 71 (70%) | 194 (72%) | 2 (50%) |
| Yes | 108 (29%) | 30 (30%) | 76 (28%) | 2 (50%) |
| History of treatment for precancer |  |  |  |  |
| No | 368 (98%) | 99 (98%) | 265 (98%) | 4 (100%) |
| Yes | 7 (2%) | 2 (2%) | 5 (2%) | 0 (0%) |

Data are n= number of participants (%). ART= antiretroviral therapy. CD4= cluster of differentiation 4. HIV RNA= human immunodeficiency virus ribonucleic acid. hrHPV= high-risk human papillomavirus. IQR= inter-quartile range. VIA= visual inspection of the uterine cervix after application of 3–5% acetic. CIN2+= Cervical intra-epithelial neoplasia grade two and above. <CIN2= Cervical intra-epithelial neoplasia grade one and below. \* =income in 3 months before enrolment. copies/mL= copies per millilitre. cells/mm3= cell per cubic millimetre.

### Disease spectrum

A consensus diagnosis of CIN2+ was made in 101 of 371 women with valid histology results (27.2%), of which 44 were CIN2, 56 were CIN3, and one was invasive cancer. The pathologists’ agreement for determining CIN2+/HSIL was 71% (κ=0.37) at baseline and 82% (κ=0.46) at follow-up. Despite efforts to link all women with CIN2+ to care (S7), only 64/101 received treatment. Of these, 50 did not attend follow-up, four had positive histology at follow-up, and 10 had negative histology results at follow-up. Prevalence of hrHPV was 43.5% (163/371) and *T. vaginalis* 19% (70/371) (S8).

### Stand-alone screening test-accuracy

Of 101 women with CIN2+, 23 (22.8%) had a negative result on all three screening tests (Table 2). The standalone test with the highest point estimate for sensitivity was hrHPV testing (67.3%, 95% CI: 57.7–75.7) (Figure 2, Table of results in S9). Specificity was 65.3% (95% CI: 59.4–70.7). Women with CIN2+ were almost four times more likely to test positive for hrHPV than those without (DOR hrHPV 3.9, 95% CI: 2.4–6.3). Using the Swede score, the AUC for Gynocular was 0.69 (95% CI: 0.63–0.75) (S10). When dichotomised using the Youden index (Swede score 3), the test had a sensitivity of 51.5% (95% CI: 41.9–61.0), a specificity of 80.0% (95% CI: 74.8–84.3), and DOR of 4.25 (95% CI: 2.6–6.9, Figure 2, S9). When using the Swede score 1 (threshold yielding sensitivity ≥ 90%), we reached a sensitivity of 97.0% (95% CI: 92.0–99.0) with a specificity of 3.3% (95% CI: 1.8–6.2). When using the Swede score 6 (threshold yielding specificity ≥ 90%), specificity reached 94.1% (95% CI: 90.6–96.3) with a sensitivity of 29.7% (95% CI: 21.7–39.2). VIA had the lowest sensitivity (22.8%, 95% CI: 15.7–31.9) and highest specificity (92.6%, 95% CI: 88.8–95.2), with a DOR of 3.7 (95% CI: 1.9–7.1).

**Table 2.**
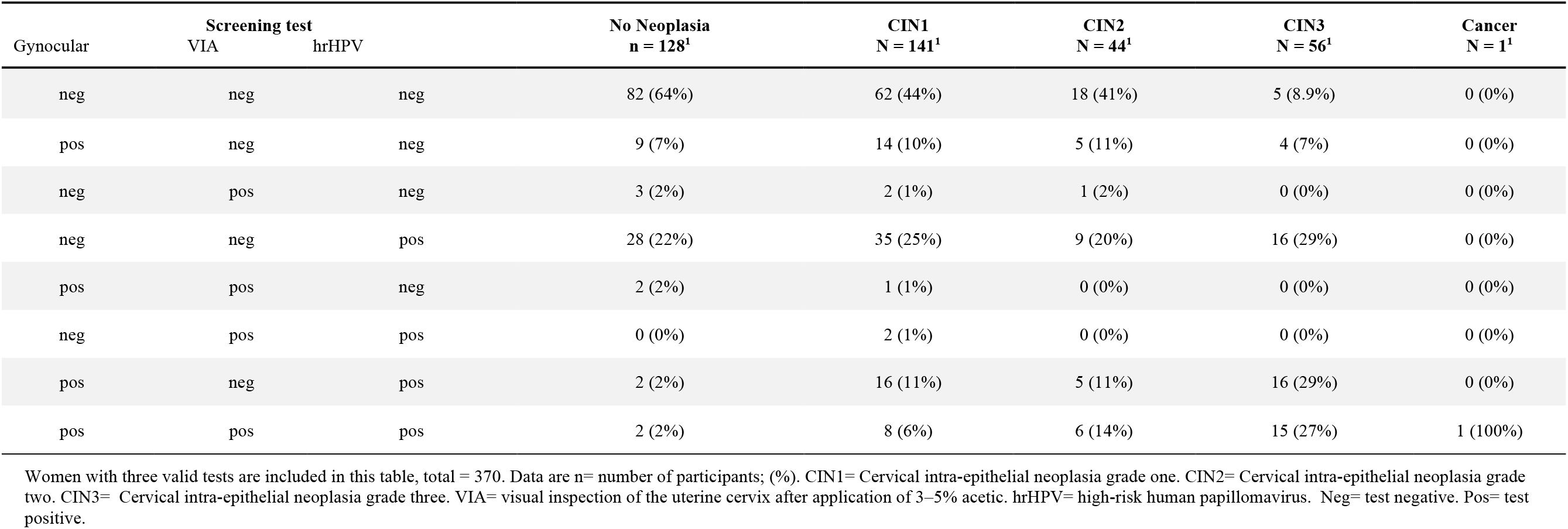
Tests results and CIN status.

| Gynocular | Screening test |  | No Neoplasia<br>n = 128<sup>1</sup> | CIN1<br>N = 141<sup>1</sup> | CIN2<br>N = 44<sup>1</sup> | CIN3<br>N = 56<sup>1</sup> | Cancer<br>N = 1<sup>1</sup> |
| --- | --- | --- | --- | --- | --- | --- | --- |
|  | VIA | hrHPV |  |  |  |  |  |
| neg | neg | neg | 82 (64%) | 62 (44%) | 18 (41%) | 5 (8.9%) | 0 (0%) |
| pos | neg | neg | 9 (7%) | 14 (10%) | 5 (11%) | 4 (7%) | 0 (0%) |
| neg | pos | neg | 3 (2%) | 2 (1%) | 1 (2%) | 0 (0%) | 0 (0%) |
| neg | neg | pos | 28 (22%) | 35 (25%) | 9 (20%) | 16 (29%) | 0 (0%) |
| pos | pos | neg | 2 (2%) | 1 (1%) | 0 (0%) | 0 (0%) | 0 (0%) |
| neg | pos | pos | 0 (0%) | 2 (1%) | 0 (0%) | 0 (0%) | 0 (0%) |
| pos | neg | pos | 2 (2%) | 16 (11%) | 5 (11%) | 16 (29%) | 0 (0%) |
| pos | pos | pos | 2 (2%) | 8 (6%) | 6 (14%) | 15 (27%) | 1 (100%) |

Women with three valid tests are included in this table, total = 370. Data are n= number of participants; (%). CIN1= Cervical intra-epithelial neoplasia grade one. CIN2= Cervical intra-epithelial neoplasia grade two. CIN3= Cervical intra-epithelial neoplasia grade three. VIA= visual inspection of the uterine cervix after application of 3–5% acetic. hrHPV= high-risk human papillomavirus. Neg= test negative. Pos= test positive.

**Figure 2.**
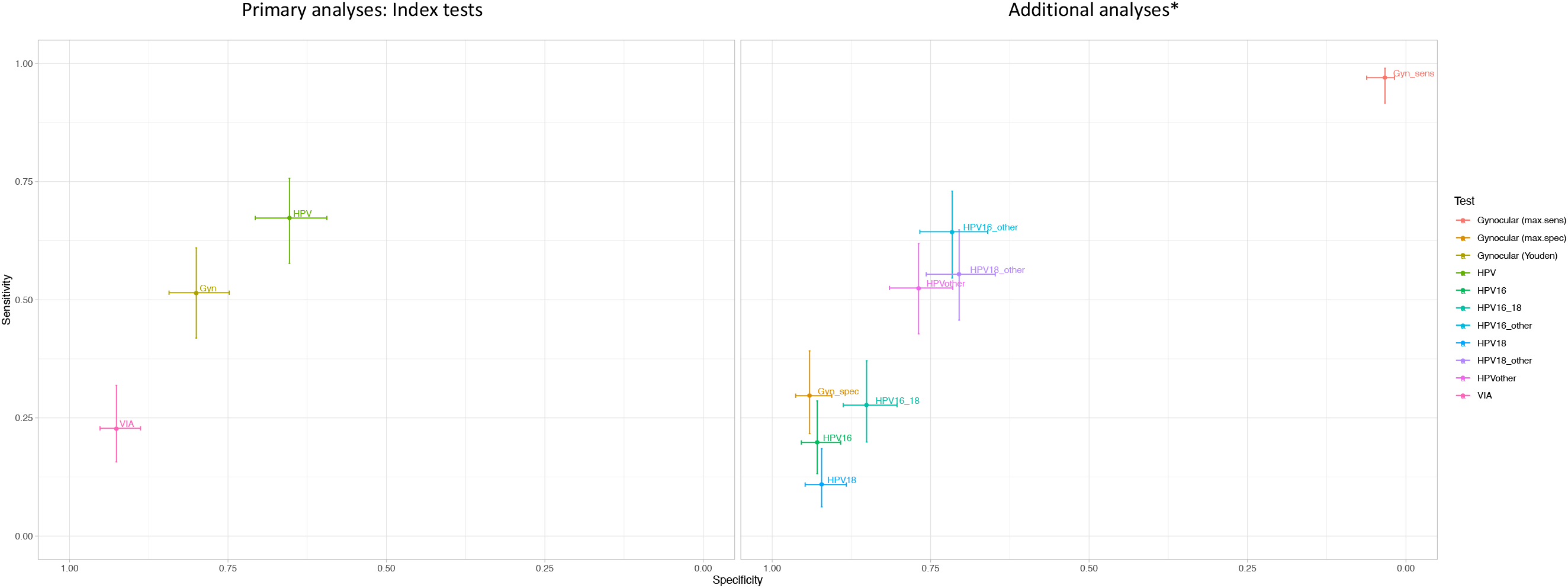
Sensitivity and specificity of single test screening strategies for prevalent CIN2+. *= Secondary analysis (Gynocular™) Sensitivity analysis (HPV subtypes). Gyn= Gynocular. Max.spec= using a threshold that maximises specificity. Max.sens= using a threshold that maximises sensitivity. HPV16= human papillomavirus subtype 16. HPV18= human papillomavirus subtypes 18 and 45. HPVother= human papillomavirus other high-risk subtypes pooled -31, 33, 35, 39, 51, 52, 56, 58, 59, 66, and 68. hrHPV= high-risk human papillomavirus. VIA= visual inspection of the uterine cervix after application of 3–5% acetic.

### Sensitivity analyses

We did not detect a strong training effect (S11a). Test-accuracy measures were similar whether or not participants stopped the study because of the COVID-19 pandemic (S11b), and results replacing missing and indeterminate test results and reference standards did not substantially affect estimates of accuracy (S11c). Using different categories of HPV subtypes showed similar results, with the best combination being HPV16 with “other” (sensitivity 64.4%, 95% CI: 54.6–73.0, specificity 71.6%, 95% CI: 66.0–76.7 and DOR 4.6, 95% CI: 2.8–7.4, Table 2). In the hypothetical scenario where biopsies were taken only from visible lesions (n=106), sensitivity increased for all tests, and specificity remained at similar levels (S11d). For hrHPV, sensitivity was 85.7% (95% CI: 73.3–92.9), specificity was 62.7% (95% CI: 57.3, 67.8), and DOR was 10.1 (95% CI: 4.4–23.2). Sensitivity of Gynocular increased to 93.9% (95% CI: 83.5–97.9), specificity to 81.5% (95% CI 76.9–85.3), and DOR to 67.5 (95% CI: 20.3–22.4). The sensitivity for VIA increased to 44.9% (95% CI: 31.9–58.7), specificity to 93.5% (95% CI: 90.3–95.7), and DOR to 11.8 (95% CI: 5.75–24.1).

### Two tests in combination

When we examined combinations of two tests with positive results, we found the specificities improved to above 90% for all test combinations but found a higher proportion of false negatives than when using single screening tests. Combining hrHPV followed by Gynocular yielded the most favourable balance of sensitivity and specificity (sensitivity 42.6%, 95% CI: 33.4–52.3), specificity 90.0% (95% CI: 85.3–92.7) (Table 3, S12). Other analyses of test combinations are reported in S13.

**Table 3:**
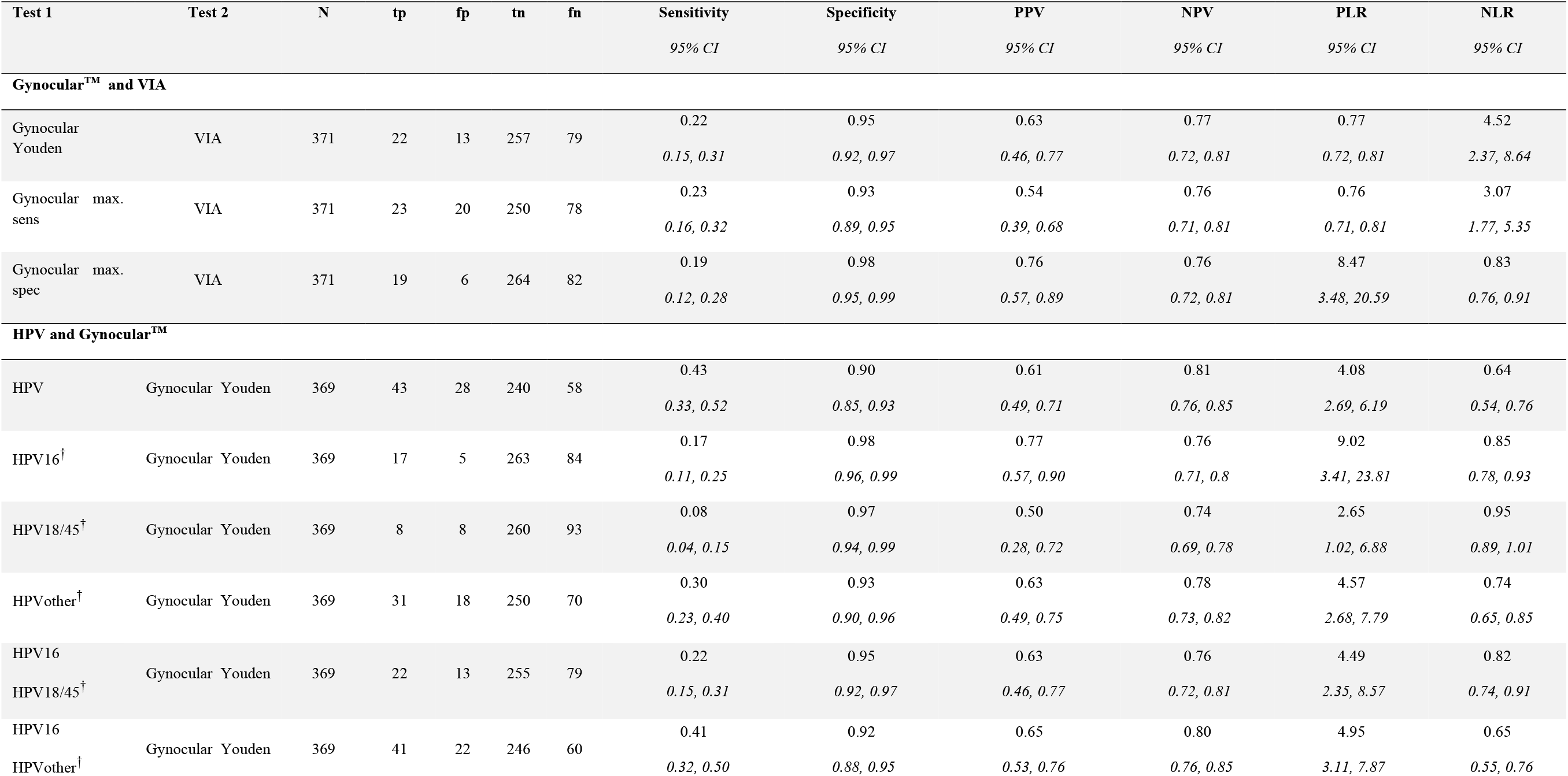
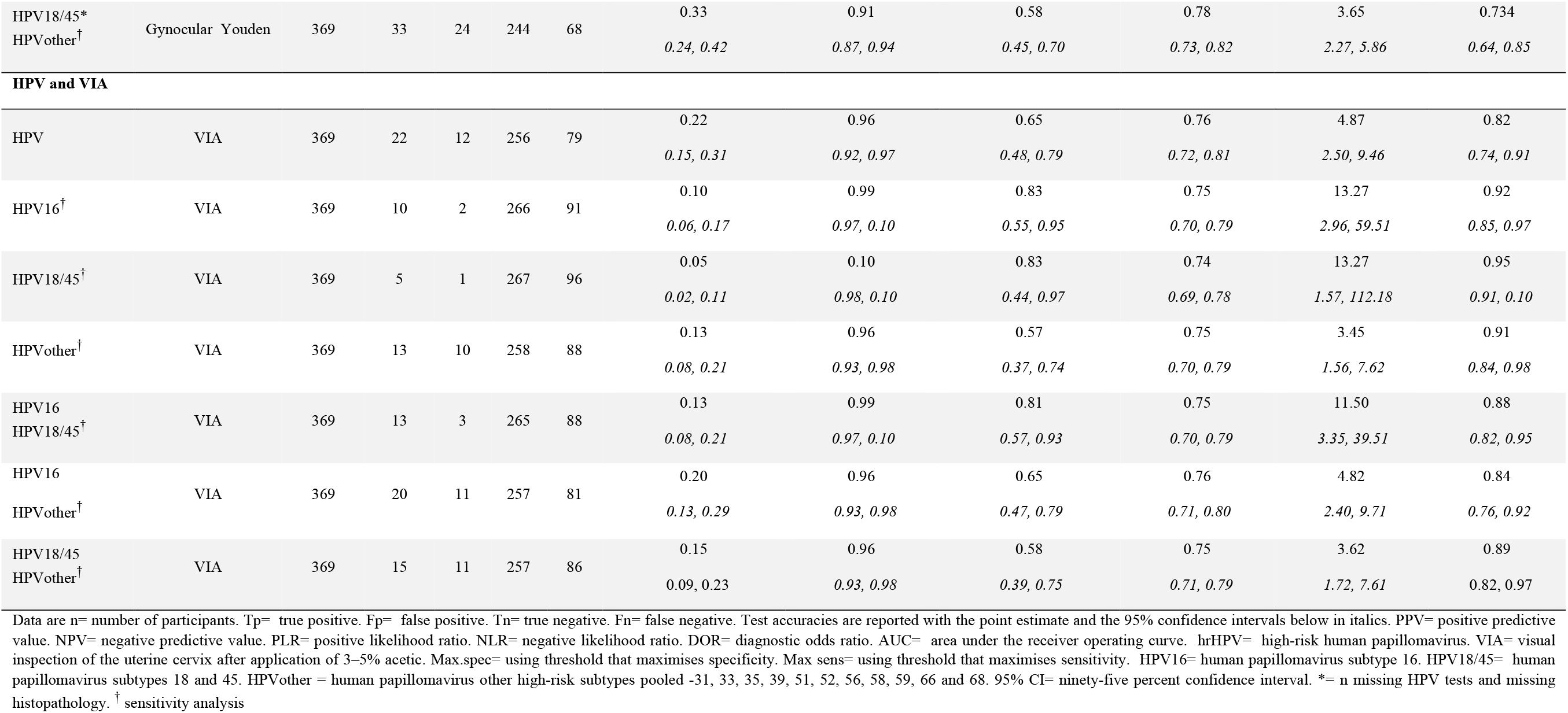
Diagnostic accuracy of tests in combination and their precision.

### Subgroup analyses

In a subgroup analysis, we found no clear differences in sensitivity and specificity according to age, menopause, education, contraception, parity, *T. vaginalis* result, ART status, HIV RNA viral load, CD4 cell count, and previous treatment for pre-cancerous disease (S14), and we did not detect effect modification by patient characteristics on the association between diagnostic test and disease status (S15).

## Discussion

We found a high prevalence of CIN2+ pre-cancerous lesions and hrHPV in WLHIV, almost all of whom were on ART. Stand-alone hrHPV, Gynocular, and VIA testing missed almost a quarter of pre-cancerous disease. Among visual screening tests, the Gynocular performed better than VIA. Combining tests did not improve test-accuracy measures. In a sensitivity analysis in which only CIN2+ detected from visible lesions was used as the reference standard, all accuracy measures improved.

The study has several strengths. First, we tested a novel magnification device (Gynocular) among WLHIV with limited access to conventional colposcopy. Second, the index tests and reference standards were relevant to the context and performed by local experts. Third, we optimised the study methods with several strategies. Local and international experts contributed to protocol development and training staff. A data safety and monitoring board provided oversight.^9^ All women received the reference standard, preventing partial verification biases. We reduced detection bias by obtaining two to four biopsies from each woman and considering the presence of disease at two timepoints six months apart. We used objective measures of HIV severity and concurrent *T. vaginalis* to examine associations between co-existing conditions and test performance.^15,16^ We safeguarded blinding of screening tests and the reference standard. Furthermore, p16 immunostaining was used to determine HSIL objectively.^11,17^ Because screening results often include indeterminate and missing results, we included a sensitivity analysis to understand the impact of these on test accuracy.

We acknowledge the limitations of our study methods. First, we used an index test (Gynocular) to guide biopsy samples for the reference standard. However, partial verification bias was avoided because all women received multiple biopsies irrespective of whether a lesion was seen. Second, the COVID-19 pandemic interrupted follow-up, and only 104 (28%) women had a second reference test by the time the study had to close. We found five additional cases of CIN2+ among these, presumably missed at baseline. Were we able to complete follow-up on all women, disease prevalence may have been higher, affecting the predictive values of the tests performed at baseline.^22^ Third, whilst we considered six months a short enough interval for the second reference standard test to detect missed disease, a 12-week timeframe has also been used.^7^ Fourth, we used GeneXpert as the hrHPV testing platform, but an additional laboratory-based method would have enhanced quality control. Fifth, the study assessed VIA, but many sites in SSA use an amended method, including cervicography.^23,24^ The results of this study are therefore not applicable to the Cervical Cancer Prevention Program in Zambia.

The sensitivity of testing for hrHPV was lower in our study than in many others.^6,7^ In contrast to many previous studies, we took four biopsies from women with no visible lesions and repeated testing six months later to avoid partial verification bias when only acetowhite lesions are sampled. We found the sensitivity of hrHPV was 65.3% (95% CI: 59.4–70.7) when biopsies were obtained from all women and 85.7% (95% CI: 73.3–92.9) if only biopsies from visible lesions were considered. Kelly *et al*.’s systematic review of cervical cancer screening strategies among WLHIV in studies published up to July 2022 found that the sensitivity of VIA was overestimated in studies with a risk of partial verification bias.^7^ They did not, however, do a subgroup analysis stratified by the risk of verification bias for hrHPV testing. Studies in which the reference standard is obtained only from visible lesions during colposcopy^24,25^ have higher estimates of sensitivity and specificity than when all women have biopsies.^6,26,27^ We also found a prevalence of pre-cancer among WLHIV that was higher than in another Zambian study, in which CIN2+ prevalence was 16% among 200 women screened at the University Teaching Hospital in 2016.^8^ A systematic review evaluating diagnostic accuracy of cervical cancer screening strategies among WLHIV found a pooled prevalence of 12% (range 2–26%),^9^ with higher prevalence in tertiary settings where referral for abnormal cervical smear or positive HPV test suggested a high risk for CIN2+. Our reference standard methods, taking two to four biopsies at two timepoints, might have detected more CIN2+ cases than in studies taking one biopsy from the most severe cervical lesion^28,29^ or a maximum of two biopsies.^6,7^ Wentzensen *et al*. found that sensitivities for detecting CIN2+ increased from 61% (95% CI: 55–67) in a single biopsy to 86% (95% CI: 80– 90) with two biopsies to 96% (95% CI: 91–99) with three biopsies.^29^ In contrast to previous studies that calculated combined test accuracy using the denominator of women testing positive from the first test, we considered all women in our denominator so as not to miss any disease in the target population. This better emulates a real-life situation highlighting that combining tests does not improve accuracy when the sensitivity of the primary screening test is low. Ideally, a screening sequence should aim for a sensitivity of 90-95% and specificity of 85% to detect CIN3+ during one screening interval.^30^ Further research is required to reach these parameters and our study highlights the need for larger test-accuracy studies among WLHIV which minimise bias, to strengthen estimates of accuracy.

In our study, hrHPV testing, Gynocular colposcopy and VIA performed poorly as standalone screening tests among WLHIV, and 22.9% of cases were not detected by any test. Combining two tests did improve specificity but not overall accuracy when all women (and all disease) were considered in the denominator. Our findings have implications for research and cervical cancer screening policies among WLHIV if test-accuracy in this high-risk population has been overestimated. According to our sensitivity analysis, the assumption that taking biopsies from visible lesions on colposcopy is an acceptable reference standard might need reassessment. WHO recommends three-to five-year screening intervals for WLHIV, based on the assumption that suboptimal screening tests at sufficiently frequent intervals will still prevent cancer because of the long pre-cancerous phase. However, if accuracy measures informing modelling studies are over-estimated, these screening intervals might be too long. Larger studies, among WLHIV, in countries with the highest disease burden and using methods that reduce verification bias are urgently required. Our robust descriptive study results can be used in future modelling studies and randomised controlled trials of screening effectiveness, both of which are needed to determine improved strategies for cervical cancer screening among WLHIV.

## Supporting information

all supplementary material

## Data Availability

The study protocol and statistical analysis plan are available upon request to the corresponding author. They can also be accessed on ClinicalTrials.gov (ref: NCT03931083). In accordance with FAIR principles, anonymised individual participant data or aggregate data will be made available upon reasonable request to the corresponding author, and with approval from all investigators.

## Acknowledgements

Thanks to Professor Matthias Egger for his initial pivotal support, his guidance in grant applications and oversight for the study. We are grateful to Jane Matambo and Jaqueline Mutale who assisted with setting up training courses, referring gynaecologists to join the study, piloting of data collection tools and providing advice on implementation. We are grateful to Dr Srabani Mittal (in memoriam), who supported an on-site training course in Zambia during the preparatory phase of this study. We thank Dr Steve Badman and Professor Andrew Vallely for sharing their experiences with the use of the GeneXpert device. We are grateful to Serra Lem Asangbeh who contributed to training, and administrative tasks for a four-month period during the study. Thanks to Eliane Rohner for her support with early versions of our data collection forms and Kate Mwendafilumba for electronic data entry. Thanks to Dr Pierre Vassilakos, who provided guidance on histological assessment of cervical biopsies. Thanks, too, to our DSMB members: Professor Patrick Petignat (chair), Dr Sven Trelle, Dr Jake Pry, Dr Ketty Lubeya and Dr Iacopo Baussano. To our clinical team, Monica Phiri Tembo, Beatrice Chibundi, thank-you for ensuring that the women enrolled in our study feel cared for and well looked after. Last but not least, thanks to the women who participated in this study.

## Data availability

The study protocol and statistical analysis plan are available upon request to the corresponding author. They can also be accessed on ClinicalTrials.gov (ref: NCT03931083). In accordance with “FAIR principles”, anonymised individual participant data or aggregate data will be made available upon reasonable request to the corresponding author, and with approval from all investigators.

## Footnotes

### Contributions

JB, NL, PB, AM and KT designed the study with input from MM, MR, AL, and AR and obtained funding for the project. MiM, HK and KT coordinated the study and oversaw the study at the study site. JB, AM, NL, PB, and MM provided supervision. MM, MiM, HK, PB, and KT provided support to the study implementation in the study sites. TM provided support to the laboratory procedures. MR did the statistical analysis with the supervision of AL. KT wrote the manuscript draft. JB, NL, AM, MiM, MR, PB, and AR provided substantial input in the manuscript and interpretation of the data. All authors had full access to all data in the study and contributed to the interpretation of data, the revision of the manuscript, approved the final version of the manuscript and had final responsibility for the decision to submit for publication.

### Patient and public involvement

Patients and/or the public were not involved in the design, or conduct, or reporting, of this research. Disemination of results will be extended to the CCPPZ and patients who have indicated they wish to be advised of the study findings.

### Provenance and peer review

Not commissioned; externally peer reviewed.

### Author note

Where authors are identified as personnel of the International Agency for Research on Cancer/World Health Organization, the authors alone are responsible for the views expressed in this article and they do not necessarily represent the decisions, policy or views of the International Agency for Research on Cancer /World Health Organization.

## References

1. World Health Organization A. WHO guideline for screening and treatment of cervical pre-cancer lesions for cervical cancer prevention, second edition. 2021, ISBN: 9789240030824. Available at: https://www.who.int/initiatives/cervical-cancer-elimination-initiative

2. Stelzle D, Tanaka LF, Lee KK, et al. Estimates of the global burden of cervical cancer associated with HIV. Lancet Glob Health. 2021;9(2):e161–e169.

3. Castellsagué X, Diaz M, de Sanjosé S, et al. Worldwide human papillomavirus etiology of cervical adenocarcinoma and its cofactors: Implications for screening and prevention. J Natl Cancer Inst. 2006;98(5):303–15.

4. Moscicki AB, Schiffman M, Burchell A, et al. Updating the natural history of human papillomavirus and anogenital cancers. Vaccine. 2012;30(SUPPL.5).

5. Morhason-Bello IO, Odedina F, Rebbeck TR, et al. Challenges and opportunities in cancer control in Africa: a perspective from the African Organisation for Research and Training in Cancer. Lancet Oncol. 2013;14(4):e142–51.

6. Smith SK, Nwosu O, Edwards A, et al. Performance of screening tools for cervical neoplasia among women in low- and middle-income countries: A systematic review and meta-analysis. Plos Global Health. 2023;2(3):e0001598

7. Kelly H, Jaafar I, Chung M, et al. Diagnostic accuracy of cervical cancer screening strategies for high-grade cervical intraepithelial neoplasia (CIN2+/CIN3+) among women living with HIV: A systematic review and meta-analysis. eClinicalMedicine. 2022;53:101645.

8. Chibwesha CJ, Frett B, Katundu K, et al. Clinical Performance Validation of 4 Point-of-Care Cervical Cancer Screening Tests in HIV-Infected Women in Zambia. J Low Genit Tract Dis. 2016;20(3):218–23.

9. Taghavi K, Moono M, Mwanahamuntu M, et al. Screening test-accuracy to improve detection of precancerous lesions of the cervix in women living with HIV: a study protocol. BMJ Open. 2020;10(12) :e037955

10. Darragh TM, Colgan T, Cox JT, et al. The Lower Anogenital Squamous Terminology Standardization Project for HPV-Associated Lesions: background and consensus recommendations from the College of American Pathologists and the American Society for Colposcopy and Cervical Pathology. J Low Genit Tract Dis. 2012;16:205–42.

11. Ozaki S, Zen Y, Inoue M. Biomarker expression in cervical intraepithelial neoplasia: potential progression predictive factors for low-grade lesions. Hum Pathol. 2011;42:1007–12.

12. Colposcopy and treatment of cervical intraepithelial neoplasia: a beginners’ manual. 2004 ISBN-13 (Database) 978-92-832-0412-1. Available at: http://screening.iarc.fr/colpochap.php?lang=1&chap=4

13. Bowring J, Strander B, Young M, et al. The Swede Score. J Low Genit Tract Dis. 2010;14(4):301–5.

14. Kalubula M, Shen H, Khanam T. Assessment of carcinogenic and toxic substances in “Insunko” herb. Toxicol reports. 2020;7:468–74.

15. Whiting PF, Rutjes AWS, Westwood ME, et al. A systematic review classifies sources of bias and variation in diagnostic test-accuracy studies. J Clin Epidemiol. 2013;66(10):1093–104.

16. Ransohoff DF, Feinstein AR. Problems of Spectrum and Bias in Evaluating the Efficacy of Diagnostic Tests. N Engl J Med. 1978;299(17):926–30.

17. Mcgrath CJ, Garcia R, Trinh TT, et al. Role of p16 testing in cervical cancer screening among HIV-infected women. PLoS One. 2017;12(10):e0185597.

18. Leeflang MMG, Rutjes AWS, Reitsma JB, et al. Variation of a test’s sensitivity and specificity with disease prevalence. CMAJ. 2013;185(11):E537–44.

19. Parham GP, Mwanahamuntu MH, Kapambwe S, et al. Population-level scale-up of cervical cancer prevention services in a low-resource setting: development, implementation, and evaluation of the cervical cancer prevention program in Zambia. Meyers C, ed. PLoS One. 2015;10(4):e0122169.

20. Mwanahamuntu MH, Sahasrabuddhe VV, Blevins M, et al. Monitoring the performance of “screen-and-treat” cervical cancer prevention programs. Int J Gynecol Obstet. 2014;126(1):88–89.

21. Huh WK, Sideri M, Stoler M, et al. Relevance of random biopsy at the transformation zone when colposcopy is negative. Obstet Gynecol. 2014 Oct;124(4):670–678.

22. Wentzensen N, Walker JL, Gold MA, et al. Multiple biopsies and detection of cervical cancer precursors at colposcopy. J Clin Oncol. 2015;33(1):83–89.

23. World Health Organisation B. WHO prequalification of in vitro diagnostics: public report. PQDx 0268-070-00 WHO PQ public report December 2017/version 3.0, 2017. Available: https://www.who.int/diagnostics_laboratory/evaluations/pq-list/hiv-vrl/171221_final_pq_report_pqdx_0268_070_00.pdf?ua=1

24. Kuhn L, Saidu R, Boa R, et al. Clinical evaluation of modifications to a human papillomavirus assay to optimise its utility for cervical cancer screening in low-resource settings: a diagnostic accuracy study. Lancet Glob Health. 2020 Feb;8(2):e296–e304.

25. Firnhaber C, Mayisela N, Mao L, et al. Validation of cervical cancer screening methods in HIV positive women from Johannesburg South Africa. Plos one. 2013;8(1):e53494

26. Kremer WW, van Zummeren M, Breytenbach E, et al. The use of molecular markers for cervical screening of women living with HIV in South Africa. AIDS. 2019 Nov 1;33(13):2035–2042.

27. Chung MH, McKenzie KP, De Vuyst H, et al. Comparing Papanicolau smear, visual inspection with acetic acid and human papillomavirus cervical cancer screening methods among HIV-positive women by immune status and antiretroviral therapy. AIDS. 2013;27(18):2909–19.

28. Wentzensen N, Walker J, Smith K, et al. A prospective study of risk-based colposcopy demonstrates improved detection of cervical precancers. Am J Obstet Gynecol. 2018 Jun;218(6):604.e1-604.e8.

29. Baasland I, Hagen B, Vogt C, et al. Colposcopy and additive diagnostic value of biopsies from colposcopy-negative areas to detect cervical dysplasia. Acta Obstet Gynecol Scand. 2016;95(11):1258–63.

30. Castle PE. The potential utility of HPV genotyping in screening and clinical management. J Natl Compr Canc Netw. 2008;6(1):83–95.

