## Supplementary material for "Accuracy of screening tests for cervical pre-cancer in women living with HIV in low-resource settings: a paired prospective study in Lusaka, Zambia": all supplementary material

### S1. Supplementary material 1: STARD checklist

| Section & Topic | No | Item | Line |
|---|---|---|---|
| **TITLE OR ABSTRACT** | | | |
| | 1 | Identification as a study of diagnostic accuracy using at least one measure of accuracy (such as sensitivity, specificity, predictive values, or AUC) | 3 |
| **ABSTRACT** | | | |
| | 2 | Structured summary of study design, methods, results, and conclusions (for specific guidance, see STARD for Abstracts) | |
| **INTRODUCTION** | | | |
| | 3 | Scientific and clinical background, including the intended use and clinical role of the index test | 109-117 |
| | 4 | Study objectives and hypotheses | 116-117 |
| **METHODS** | | | |
| *Study design* | 5 | Whether data collection was planned before the index test and reference standard were performed (prospective study) or after (retrospective study) | 127 |
| *Participants* | 6 | Eligibility criteria | 129-131 |
| | 7 | On what basis potentially eligible participants were identified (such as symptoms, results from previous tests, inclusion in registry) | 128 |
| | 8 | Where and when potentially eligible participants were identified (setting, location and dates) | 128 |
| | 9 | Whether participants formed a consecutive, random or convenience series | 128 |
| *Test methods* | 10a | Index test, in sufficient detail to allow replication | 164-177 and supplement |
| | 10b | Reference standard, in sufficient detail to allow replication | 143-162 |
| | 11 | Rationale for choosing the reference standard (if alternatives exist) | 143-162 |
| | 12a | Definition of and rationale for test positivity cut-offs or result categories of the index test, distinguishing pre-specified from exploratory | 178-189 |
| | 12b | Definition of and rationale for test positivity cut-offs or result categories of the reference standard, distinguishing pre-specified from exploratory | 178-189 |
| | 13a | Whether clinical information and reference standard results were available to the performers/readers of the index test | 163-177 |
| | 13b | Whether clinical information and index test results were available to the assessors of the reference standard | 153 |
| *Analysis* | 14 | Methods for estimating or comparing measures of diagnostic accuracy | 189-222 |
| | 15 | How indeterminate index test or reference standard results were handled | 219 |
| | 16 | How missing data on the index test and reference standard were handled | 219 |
| | 17 | Any analyses of variability in diagnostic accuracy, distinguishing pre-specified from exploratory | 215-222 |
| | 18 | Intended sample size and how it was determined | 191 -194 and supplement |
| **RESULTS** | | | |
| *Participants* | 19 | Flow of participants, using a diagram | 230-236 |
| | 20 | Baseline demographic and clinical characteristics of participants | 236-245 |
| | 21a | Distribution of severity of disease in those with the target condition | 250-256 |
| | 21b | Distribution of alternative diagnoses in those without the target condition | 252-253 and supplement |

| | | | |
|---|---|---|---|
| | 22 | Time interval and any clinical interventions between index test and reference standard | 236 |
| *Test results* | 23 | Cross tabulation of the index test results (or their distribution) by the results of the reference standard | Table 3 |
| | 24 | Estimates of diagnostic accuracy and their precision (such as 95% confidence intervals) | 258-270 table 3 and Figure 4 |
| | 25 | Any adverse events from performing the index test or the reference standard | 233 |
| **DISCUSSION** | | | |
| | 26 | Study limitations, including sources of potential bias, statistical uncertainty, and generalisability | 313-323 |
| | 27 | Implications for practice, including the intended use and clinical role of the index test | 352-362 |
| **OTHER INFORMATION** | | | |
| | 28 | Registration number and name of registry | 56 |
| | 29 | Where the full study protocol can be accessed | 56 |
| | 30 | Sources of funding and other support; role of funders | 53 |

### S2. Supplementary material 2: Description of procedures

The VIA examination included insertion of a speculum, visualisation of the vagina, vulva and cervix, assessment with the naked eye after application of normal saline, and further assessment after application of 5% acetic acid for one minute. The nurse recorded the findings as normal, abnormal, or suspicious of cancer. As per local guidelines, VIA nurses categorised indeterminate findings as abnormal.

The HPV sample was collected during the first speculum examination that participants received, and before the VIA examination. The hrHPV testing was done using a single-use cervical cytobroom provided by GeneXpert™ and placed into ThinPrep PreservCyt (Cepheid, Sunnyvale, CA) immediately after collection. ined Nurses were trained to insert the central bristles of the cervical cytobroom into the endocervical canal and to allow the shorter bristles to remain in full contact with the ectocervix, rotating the broom five times in a clockwise direction to collect cells from the transformation zone. The broom was then rinsed into the PreservCyt® solution without delay and separated into the vial without touching surrounding sites.

The nurse performing colposcopy and obtaining biopsies received training from a gynaecologist based at the International Agency for Research on Cancer (IARC) and a local senior gynaecologist. The Gynocular™ examination included a speculum examination and visualisation of the vagina, vulva and cervix, assessment of the cervix at low and high magnification (>6x), examination of cervical vessel patterns using the red-free mode (or green filter), application of 5% acetic acid for one minute, and cervical assessment following application of Lugol's iodine.

The Swede score describes the Gynocular™ examination by scoring the following domains: vessels, margins or surface, acetic acid uptake, iodine staining, and lesion size. A score of zero and two is given to each domain based on the severity of the findings, and summed to a total score between zero (best) and ten (worst). Ablative treatment was offered if the lesion boundaries were fully visible, covered less than 75% of the ectocervix, did not extend into the endocervical canal, and was covered by the cryotherapy/ thermoablation tip. For larger lesions, women were offered loop electrosurgical excision procedure.

#### Training

We trained our study team on the study protocol, data-entry, and clinical procedures twice prior to study commencement and once during the study period. Clinical training sessions were led by a local expert gynaecologist as well as IARC trainers. We taught all gynaecological procedures on mannequins first and then on women at the cervical cancer screening clinic at Kanyama hospital under supervision. Cepheid company representatives in Lusaka set up and calibrated the GeneXpert™ device yearly. They provided two training sessions on obtaining and processing hrHPV swabs; one before the study commencement and one after a year.

#### Treatment

All women with VIA-positive findings, or CIN2+ on histology, were offered treatment with cryotherapy, thermoablation, or loop electrosurgical excision procedure (LEEP) as clinically indicated. Women with histopathologically confirmed cervical cancer were referred to the University Teaching Hospital in Lusaka for treatment.

**S3. Supplementary material 3: The Gynocular$^{TM}$ (Gynius Plus AB, Sweden)**

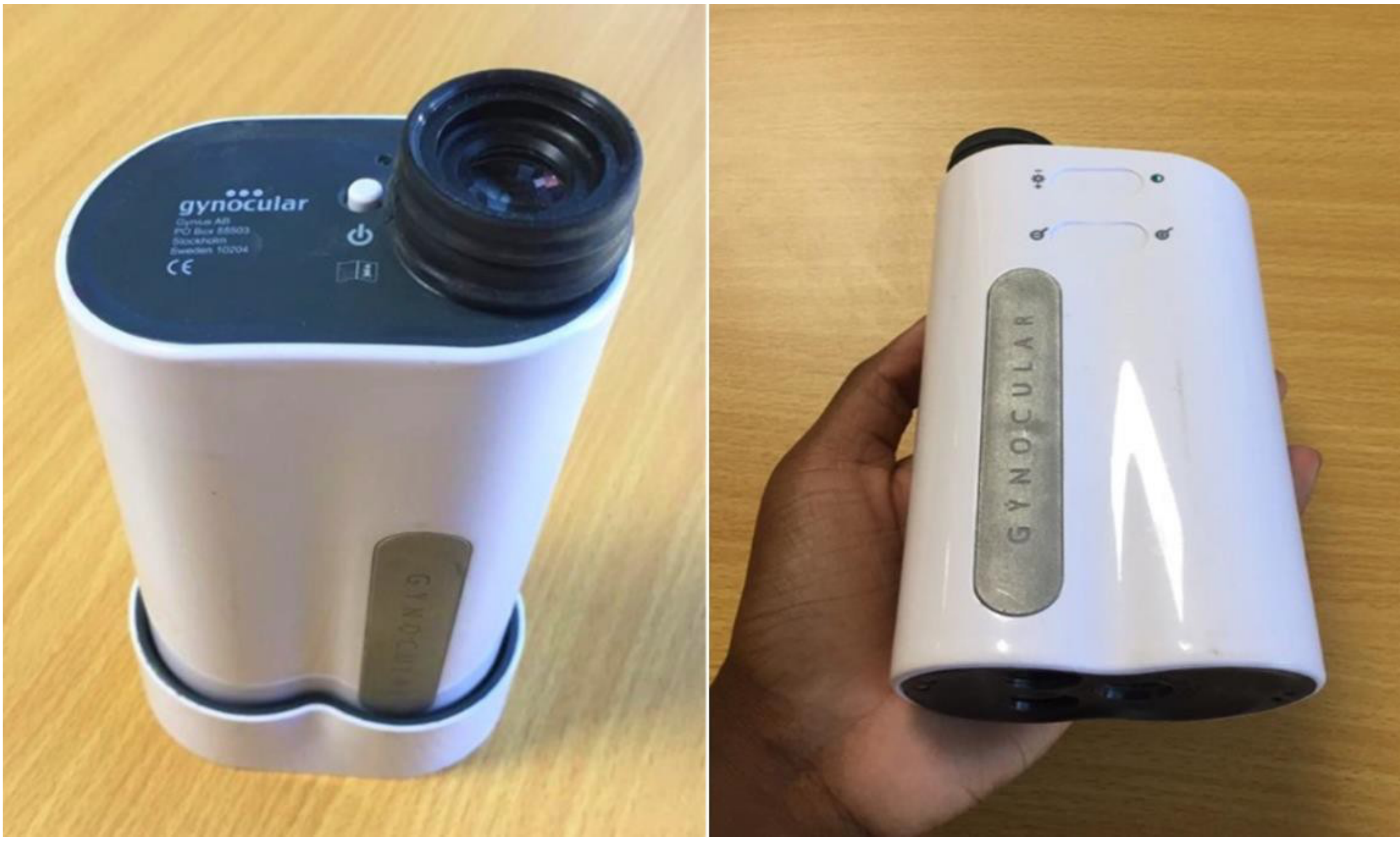

This photograph was taken at the Centre for Infectious Disease Research Zambia headquarters on 27 April 2020 for this manuscript.

### S4. Supplementary material 4: Swede score definition

| Swede score | 0 | 1 | 2 |
|---|---|---|---|
| **Aceto uptake** | Zero or transparent | Shady milky (not transparent not opaque) | Distinct, opaque white |
| **Margins/surface** | Diffuse | Sharp but irregular, jagged, "geographical" satellites | Sharp and even, difference in surface level including "cuffing" |
| **Vessels** | Fine, regular | Absent | Coarse or atypical |
| **Lesion size** | <5mm | 5-15mm or 2 quadrants | >15mm or 3-4 quadrants or endocervically undefined |
| **Iodine staining** | Brown | Faintly or patchy yellow | Distinct yellow |
| | | | **TOTAL SCORE** |

Source: Bowring J, Strander B, Young M, Evans H, Walker P. The Swede score: evaluation of a scoring system designed to improve the predictive value of colposcopy. J Low Genit Tract Dis. 2010 Oct;14(4):301-5. doi: 10.1097/LGT.0b013e3181d77756. PMID: 20885156.

### S5. Supplementary material 5: Sample-size calculation

This screening-test accuracy study requires 350 participants to estimate the sensitivity and specificity of Gynocular™, HR-HPV, and VIA for CIN2+ lesions with the precision detailed in **Table S5**. Screening accuracy measures will be estimated with 95% Wilson confidence intervals with no formal hypothesis testing between modalities.

We planned to enrol 450 women to obtain data from at least 350 patients for statistical analyses. We expected the prevalence of CIN2+ in WLHIV in Zambia to be 16–20%.[1-3] We expected disease prevalence to be lower than in previous years. Increases in the number of women receiving ART, and commencing treatment at higher CD4 cell counts,[4] may lead to a decline in HPV prevalence.[5] Higher rates of voluntary male circumcision in male partners may also contribute to lower levels of HPV infection.[5] We also estimated that there may be up to 10% loss to follow-up and up to 10% of tests that are not analysable or interpretable. We implemented rigorous data collection methods, such as patient demographics, clinical history, and test results, to avoid missing data.

**Table S5:** Sensitivity and specificity table, N=350. The expected 95% Wilson confidence interval (%) for sensitivity and specificity with varying prevalence.

| | **Expected 95% confidence intervals for sensitivity (%)**<br>***Expected 95% confidence intervals for specificity (%)*** | | | | | | | | | |
|---|---|---|---|---|---|---|---|---|---|---|
| **Expected prevalence (%)** | **50** | **55** | **60** | **65** | **70** | **75** | **80** | **85** | **90** | **95** |
| **14** | 37.5–64.4<br>*44.6–55.8* | 41.3–68.1<br>*49.5–60.7* | 45.2–71.8<br>*54.5–65.5* | 51.3–77.1<br>*59.6–70.3* | 55.5–80.5<br>*64.7–75.0* | 61.9–85.4<br>*69.9–79.6* | 66.4–88.5<br>*75.2–84.2* | 73.3–92.9<br>*80.6–88.6* | 78.2–95.6<br>*86.1–92.9* | 86.3–98.9<br>*91.9–97.0* |
| **16** | 37.3–62.7<br>*44.3–55.7* | 42.4–67.6<br>*49.4–60.7* | 47.6–72.4<br>*54.2–65.3* | 51.2–75.5<br>*59.4–70.2* | 56.7–80.1<br>*64.6–75.0* | 62.3–84.5<br>*69.9–79.8* | 68.2–88.7<br>*75.0–84.1* | 74.3–92.6<br>*80.5–88.7* | 78.5–95.0<br>*86.2–93.0* | 85.4–98.2<br>*91.8–96.9* |
| **18** | 38.8–62.7<br>*44.4–55.9* | 43.3–67.2<br>*49.3–60.7* | 48.0–71.5<br>*54.2–65.4* | 52.8–75.7<br>*59.5–70.4* | 57.6–79.8<br>*64.5–75.0* | 62.7–83.7<br>*69.6–79.6* | 67.8–87.5<br>*75.1–84.3* | 75.0–92.3<br>*80.4–88.7* | 80.7–95.6<br>*85.9–92.9* | 86.9–98.4<br>*92.0–97.1* |
| **20** | 38.6–61.4<br>*44.2–55.8* | 44.1–66.8<br>*49.1–60.7* | 48.3–70.7<br>*54.2–65.6* | 54.0–75.8<br>*59.2–70.3* | 58.5–79.5<br>*64.4–75.1* | 64.5–84.2<br>*69.6–79.7* | 69.2–87.7<br>*74.9–84.3* | 75.7–92.1<br>*80.3–88.7* | 80.8–95.1<br>*85.9–93.0* | 88.1–98.5<br>*91.8–97.0* |
| **22** | 39.7–61.5<br>*44.3–56.1* | 43.5–65.2<br>*49.0–60.7* | 48.6–70.0<br>*54.2–65.7* | 53.8–74.7<br>*59.0–70.3* | 59.2–79.2<br>*64.3–75.1* | 64.6–83.6<br>*69.6–79.8* | 70.3–87.8<br>*74.7–84.2* | 74.7–90.9<br>*80.3–88.7* | 80.8–94.6<br>*86.0–93.1* | 87.4–98.0<br>*91.6–96.9* |

1. Chibwesha CJ, Frett B, Katundu K, et al. Clinical performance validation of 4 point-of-care cervical cancer screening tests in HIV-infected women in Zambia. J Low Genit Tract Dis 2016;20:218–23.doi:10.1097/LGT.0000000000000206pmid:http://www.ncbi.nlm.nih.gov/pubmed/27030883
2. Bateman AC, Katundu K, Mwanahamuntu MH, et al. The burden of cervical pre-cancer and cancer in HIV positive women in Zambia: a modeling study. BMC Cancer 2015;15:541. doi:10.1186/s12885-015-1558-5pmid:http://www.ncbi.nlm.nih.gov/pubmed/262059
3. Parham GP, Sahasrabuddhe VV, Mwanahamuntu MH, et al. Prevalence and predictors of squamous intraepithelial lesions of the cervix in HIV-infected women in Lusaka, Zambia. Gynecol Oncol 2006;103:1017–22.doi:10.1016/j.ygyno.2006.06.015pmid:http://www.ncbi.nlm.nih.gov/pubmed/16875716
4. World Health Organization. Department of HIV/AIDS, guideline on when to start antiretroviral therapy and on pre-exposure prophylaxis for HIV, 2015.
5. Hall MT, Smith MA, Simms KT, et al. The past, present and future impact of HIV prevention and control on HPV and cervical disease in Tanzania: a modelling study. PLoS One 2020;15:e0231388.

**S6. Supplementary material 6: Baseline characteristics of all women aged between 18 and 65 years seen at Kanyama clinic over the recruitment period of the study**

| Characteristic | N = 10,718 |
|---|---|
| Highest educational level | |
| None | 546 (6.2%) |
| Finished primary | 3,524 (40%) |
| Started secondary | 44 (0.5%) |
| Finished secondary | 4,491 (51%) |
| College/University | 156 (1.8%) |
| Unknown | 1,957 |
| Marital status | |
| In a relationship/Married | 5,864 (66%) |
| Separated/Divorced | 1,256 (14%) |
| Single | 902 (10%) |
| Widowed | 923 (10%) |
| Unknown | 1,773 |
| Age | |
| Median (IQR) | 35 (28, 43) |
| Income [Kwacha/month] | |
| Greater than K3000 | 54 (2.1%) |
| K1000 - K1499 | 678 (27%) |
| K1500 - K1999 | 231 (9.0%) |
| K2000 - K2999 | 201 (7.9%) |
| K500 - K999 | 935 (37%) |
| Less than K500 | 454 (18%) |
| Unknown | 8,165 |
| Gravidity | |
| Median (IQR) | 1.00 (1.00, 3.00) |
| Unknown | 9,358 |
| Parity | |
| Median (IQR) | 0.00 (0.00, 1.00) |
| Unknown | 9,358 |
| Contraception use | |
| Yes | 2,596* |
| Unknown | 8,122 |
| HIV RINA Viral load | |
| < 50 copies/ml | 5,503 (73%) |
| [50,1000) | 1,386 (18%) |
| [1000,10000) | 226 (3.0%) |
| 10000 and more | 456 (6.0%) |
| Unknown | 3,147 |

*percentage not provided as it would be misleading, some of the women categorized with contraception status unknow are likely not using contraception, however it was not possible to differentiate the number from the raw data source. Categories have been aligned as much as possible with baseline characteristics reported in our study. HIV RNA, human immunodeficiency virus ribonucleic acid; hrHPV, high-risk human papillomavirus; IQR, inter-quartile range; n, number of participants; VIA, visual inspection of the uterine cervix after application of 3–5% acetic; CIN2+, Cervical intra-epithelial neoplasia grade two and above; <CIN2, Cervical intra-epithelial neoplasia grade one and below; copies/mL: copies per millilitre; cells/mm3: cell per cubic millimetre.

### S7. Supplementary material 7: Measures taken to link women to treatment

1) Women were called, and a peer educator was sent into the community to encourage women to attend treatment appointments. Women were offered repeat appointments until they attended. On average, women were offered 3 appointments.
2) Additional travel money was provided for women to attend treatments.
3) Gynaecologist was hired and available daily at the screening clinic to perform LEEPs
4) An additional gynaecologist came to treat study patients once per week

### S8. Supplementary material 8: Distribution of alternative diagnoses in those without the target condition

| | **Negative** | | **Positive** | | |
|---|---|---|---|---|---|
| | Normal tissue (n=129) | CIN1 (n=141) | CIN2 (n=44) | CIN3 (n=56) | Cancer (n=1) |
| **T. vaginalis** | | | | | |
| *Negative* | *106 (82.2%)* | *117 (83.0%)* | *34 (77.3%)* | *45 (80.4%)* | *0 (0%)* |
| *Positive* | *23 (17.8%)* | *24 (17.0%)* | *10 (22.7%)* | *11 (19.6%)* | *1 (100%)* |
| **HPV** | | | | | |
| *Negative* | *96 (74.4%)* | *79 (56.0%)* | *24 (54.5%)* | *9 (16.1%)* | *0 (0%)* |
| *Positive* | *32 (24.8%)* | *61 (43.3%)* | *20 (45.5%)* | *47 (83.9%)* | *1 (100%)* |
| *Undetermined* | *1 (0.8%)* | *1 (0.7%)* | *0 (0%)* | *0 (0%)* | *0 (0%)* |

### S9. Supplementary material 9: Estimates of diagnostic accuracy and their precision

| Test | n | tp | fp | tn | fn | Sensitivity %<br>*95% CI* | Specificity %<br>*95% CI* | PPV %<br>*95% CI* | NPV %<br>*95% CI* | PLR<br>*95% CI* | NLR<br>*95% CI* | DOR<br>*95% CI* | AUC |
|---|---|---|---|---|---|---|---|---|---|---|---|---|---|
| **Primary analysis** | | | | | | | | | | | | | |
| Gynocular<br>(Youden) | 371 | 52 | 54 | 216 | 49 | 51.5<br>*41.9 - 61.0* | 80.0<br>*74.8 - 84.3* | 49.1<br>*39.7 - 58.4* | 81.5<br>*76.4 - 85.7* | 2.6<br>*1.9 - 3.5* | 0.6<br>*0.5, 0.7* | *4.2*<br>*2.6 - 6.9* | 0.69<br>0.63 - 0.75 |
| hrHPV | 369* | 68 | 93 | 175 | 33 | 67.3<br>*57.7 - 75.7* | 65.3<br>*59.4 - 70.7* | 41.6<br>*34.3 - 49.3* | 84.1<br>*78.6 - 88.5* | 1.9<br>*1.6 - 2.4* | 0.5<br>*0.4 - 0.7* | *3.9*<br>*2.4 - 6.3* | |
| VIA | 371 | 23 | 20 | 250 | 78 | 22.8<br>*15.7 - 31.9* | 92.6<br>*88.8 - 95.2* | 51.2<br>*36.8 - 65.4* | 76.2<br>*71.3 - 80.5* | 3.1<br>*1.8 - 5.3* | 0.8<br>*0.7 - 0.9* | *3.7*<br>*1.9 -7.1* | |
| **Additional analyses** | | | | | | | | | | | | | |
| Gynocular<br>(max.spec) | 371 | 30 | 16 | 254 | 71 | 29.7<br>*21.7 - 39.2* | 94.1<br>*90.6 - 96.3* | 65.2<br>*50.8 - 77.3* | 78.2<br>*73.3 - 82.3* | 1.0<br>*1.0 - 1.1* | 0.9<br>*0.3 - 3.2* | 6.7<br>*3.46 - 13.0* | 0.69<br>*0.63 - 0.75* |
| HPV16<br>HPVother† | 369* | 65 | 76 | 192 | 36 | 64.4<br>*54.6 - 73.0* | 71.6<br>*66.0 - 76.7* | 46.1<br>*38.1 - 54.3* | 84.2<br>*78.9 - 88.4* | 2.3<br>*1.8 - 2.9* | 0.5<br>*0.4 - 0.7* | 4.6<br>*2.8 - 7.4* | |
| HPVother† | 369* | 53 | 62 | 206 | 48 | 52.5<br>*42.8 - 61.9* | 76.9<br>*71.5 - 81.5* | 46.1<br>*37.3 - 55.2* | 81.1<br>*75.8 - 85.4* | 2.3<br>*1.7 - 3.0* | 0.6<br>*0.5 - 0.8* | 3.7<br>*2.3 - 5.9* | |
| HPV16† | 369* | 20 | 19 | 249 | 81 | 19.8<br>*13.2 - 28.6* | 92.9<br>*89.2 - 95.4* | 51.3<br>*36.2 - 66.1* | 75.5<br>*70.5 - 79.8* | 2.8<br>*1.6 - 5.0* | 0.9<br>*0.8 - 1.0* | 3.236<br>*1.6 - 6.4* | |
| HPV18/45<br>HPVother† | 369* | 56 | 79 | 189 | 45 | 55.4<br>*45.7 - 64.8* | 70.5<br>*64.8 - 75.7* | 41.5<br>*33.5 - 49.9* | 80.8<br>*75.2 - 85.3* | 1.8<br>*1.5 - 2.4* | 0.6<br>*0.5 - 0.8* | 3.0<br>*1.9 - 4.8* | |
| HPV16<br>HPV18/45† | 369* | 28 | 40 | 228 | 73 | 27.7<br>*19.9 - 37.1* | 85.1<br>*80.3 - 88.8* | 41.2<br>*30.3 - 53.0* | 75.7<br>*70.6 - 80.2* | 1.9<br>*1.2 - 2.8* | 0.9<br>*0.7 - 1.0* | 2.2<br>*1.3 - 3.8* | |
| HPV18/45† | 369* | 11 | 21 | 247 | 90 | 10.9<br>*6.2 - 18.5* | 92.2<br>*88.3 - 94.8* | 34.4<br>*20.4 - 51.7* | 73.3<br>*68.3 - 77.7* | 1.4<br>*0.7 - 2.8* | 1.0<br>*0.9 - 1.0* | 1.4<br>*0.7 - 3.1* | |
| Gynocular<br>(max.sens) | 371 | 98 | 261 | 9 | 3 | 97.0<br>*91.6 - 99.0* | 3.3<br>*1.8 - 6.2* | 27.3<br>*22.9 - 32.1* | 75.0<br>*46.8 - 91.1* | 1.0<br>*1.0 - 1.1* | 0.9<br>*0.2 - 3.2* | 1.1<br>*0.3 - 4.2* | 0.69<br>*0.6 - 0.8* |

Data are n= number of participants. Tp= true positive. Fp= false positive. Tn= true negative. Fn= false negative. Test accuracies are reported with the point estimate and the 95% confidence intervals below in italics. PPV= positive predictive value. NPV= negative predictive value. PLR= positive likelihood ratio. NLR= negative likelihood ratio. DOR= diagnostic odds ratio. AUC= area under the receiver operating curve. hrHPV= high-risk human papillomavirus. VIA= visual inspection of the uterine cervix after application of 3–5% acetic. Max.spec= using threshold that maximises specificity. Max sens= using threshold that maximises sensitivity. HPV16= human papillomavirus subtype 16. HPV18/45= human papillomavirus subtypes 18 and 45. HPVother = human papillomavirus other high-risk subtypes pooled -31, 33, 35, 39, 51, 52, 56, 58, 59, 66 and 68. 95% CI= ninety-five percent confidence interval. *= n missing HPV tests and missing histopathology. † sensitivity analysis

**S10. Supplementary material 10: Area under the receiver operating curve for Gynocular™**

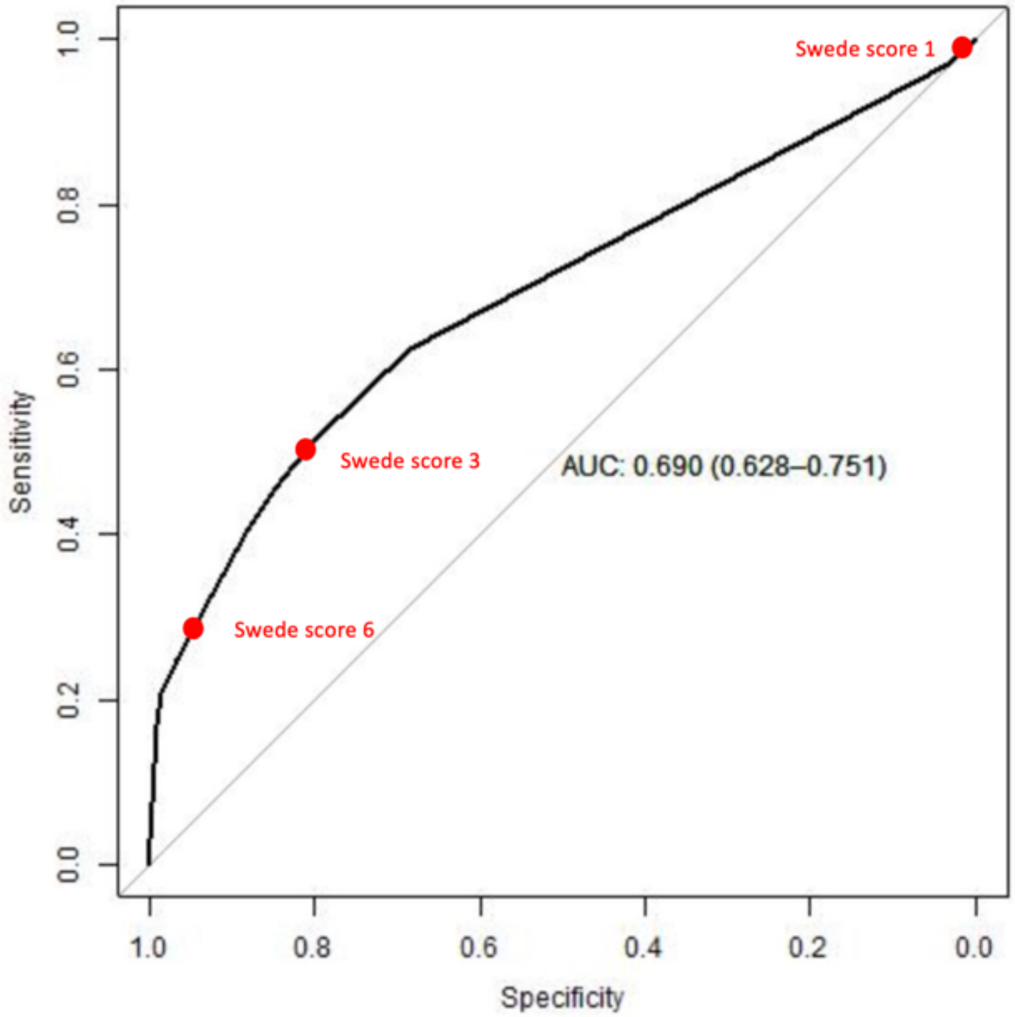

AUC= area under the curve. Swede score 6 = score of 6 when using the Swede score to determine lesion severity. Swede score 6 was associated with the maximum specificity achievable using the Gynocular™ and Swede score. Swede score 3 = score of 3 when using the Swede score to determine lesion severity. Swede score 3 achieved the Youden threshold, which maximises sensitivity and specificity. Swede score 1 = score of 1 when using the Swede score to determine lesion severity. Swede score 1 was associated with the maximum sensitivity achievable using the Gynocular™ and Swede score

### S11. Supplementary material 11: Sensitivity analyses

#### A. Training effect

Test accuracy in first 10% of patients

| | n | tp | fp | tn | fn | **Sensitivity %** *95% CI* | **Specificity %** *95% CI* | **PPV %** *95% CI* | **NPV %** *95% CI* | **PLR** *95% CI* | **NLR** *95% CI* | **DOR** *95% CI* |
|---|---|---|---|---|---|---|---|---|---|---|---|---|
| **HPV** | 36 | 6 | 8 | 21 | 1 | 0.85 [0.49, 0.97] | 0.72 [0.54, 0.85] | 0.43 [0.21, 0.67] | 0.96 [0.78, 0.99] | 3.11 [1.60, 6.03] | 0.20 [0.032, 1.228] | 15.70 [1.6 152] |
| **VIA** | 37 | 1 | 4 | 26 | 6 | 0.14 [0.03, 0.51] | 0.86 [0.70, 0.95] | 0.20 [0.04, 0.62] | 0.81 [0.65, 0.91] | 1.07 [0.14, 8.17] | 0.99 [0.709, 1.38] | 1.08 [0.10, 11.52] |
| **Gyncoluar* (youden)** | 37 | 3 | 6 | 24 | 4 | 0.42 [0.16, 0.75] | 0.80 [0.63, 0.91] | 0.33 [0.12, 0.65] | 0.86 [0.69, 0.94] | 2.14 [0.70, 6.54] | 0.71 [0.367, 1.39] | 3.00 [0.52, 17.16] |

*AUC: 0.517 [0.24, 0.793], Tp, true positive; fp, false positive; tn, true negative; fn, false negative; PPV, positive predictive value; NPV, negative predictive value; PLR, positive likelihood ratio; NLR, negative likelihood ratio; DOR, diagnostic odds ratio; AUC, area under the receiver operating curve; hrHPV, high-risk human papillomavirus; VIA , visual inspection of the uterine cervix after application of 3–5% acetic, HPV, human papillomavirus test, 95% CI, ninety-five percent confidence interval, n number of women

Test accuracy in women recruited later in the study

| | n | tp | fp | tn | fn | **Sensitivity %** *95% CI* | **Specificity %** *95% CI* | **PPV %** *95% CI* | **NPV %** *95% CI* | **PLR** *95% CI* | **NLR** *95% CI* | **DOR** *95% CI* |
|---|---|---|---|---|---|---|---|---|---|---|---|---|
| HPV | 333 | 62 | 85 | 154 | 32 | 0.66 [0.56, 0.76] | 0.64 [0.58, 0.70] | 0.42 [0.35, 0.50] | 0.83 [0.77, 0.88] | 1.86 [1.48, 2.32] | 0.528 [0.39, 0.71] | 3.51 [2.13, 5.80] |
| VIA | 334 | 22 | 16 | 224 | 72 | 0.23 [0.16, 0.33] | 0.93 [0.89, 0.96] | 0.58 [0.42, 0.72] | 0.76 [0.71, 0.80] | 3.51 [1.93, 6.39] | 0.82 [0.73, 0.92] | 4.28 [2.13, 8.59] |
| Gyncoluar* (youden) | 334 | 49 | 48 | 192 | 45 | 0.52 [0.42, 0.62] | 0.80 [0.75, 0.85] | 0.51 [0.41, 0.60] | 0.81 [0.76, 0.86] | 2.61 [1.90, 3.59] | 0.60 [0.48, 0.75] | 4.36 [2.61, 7.28] |

Tp, true positive; fp, false positive; tn, true negative; fn, false negative; PPV, positive predictive value; NPV, negative predictive value; PLR, positive likelihood ratio; NLR, negative likelihood ratio; DOR, diagnostic odds ratio; AUC, area under the receiver operating curve; hrHPV, high-risk human papillomavirus; VIA , visual inspection of the uterine cervix after application of 3–5% acetic, HPV, human papillomavirus test, 95% CI, ninety-five percent confidence interval, n number of women
AUC: 0.705 [0.642, 0.767]

### B. COVID 19

Women who stopped due to the COVID 19 pandemic

| | n | tp | fp | tn | fn | Sensitivity % *95% CI* | Specificity % *95% CI* | PPV % *95% CI* | NPV % *95% CI* | PLR *95% CI* | NLR *95% CI* | DOR *95% CI* |
|---|---|---|---|---|---|---|---|---|---|---|---|---|
| HPV | 110 | 18 | 27 | 54 | 11 | 0.62 [0.44, 0.77] | 0.667 [0.56, 0.76] | 0.40 [0.27, 0.55] | 0.83 [0.72, 0.90] | 1.86 [1.22, 2.83] | 0.57 [0.35, 0.93] | 3.27 [1.36, 7.90] |
| VIA | 111 | 7 | 6 | 76 | 22 | 0.24 [0.12, 0.42] | 0.93 [0.85, 0.97] | 0.54 [0.29, 0.77] | 0.78 [0.68, 0.85] | 3.30 [1.21, 9.01] | 0.82 [0.66, 1.01] | 4.03 [1.23, 13.24] |
| Gyncoluar* (youden) | 111 | 17 | 17 | 65 | 12 | 0.59 [0.41, 0.75] | 0.79 [0.69, 0.87] | 0.50 [0.34, 0.66] | 0.84 [0.75, 0.91] | 2.83 [1.68, 4.77] | 0.52 [0.33, 0.82] | 5.42 [2.18, 13.48] |

*AUC: 0.726 [0.613, 0.839] Tp, true positive; fp, false positive; tn, true negative; fn, false negative; PPV, positive predictive value; NPV, negative predictive value; PLR, positive likelihood ratio; NLR, negative likelihood ratio; DOR, diagnostic odds ratio; AUC, area under the receiver operating curve; hrHPV, high-risk human papillomavirus; VIA , visual inspection of the uterine cervix after application of 3–5% acetic, HPV, human papillomavirus test, 95% CI, ninety-five percent confidence interval, n number of women. NB: n=110 includes the total number of women who received followup before study stopped due to COVID and women who had to stop due to pregnancy.

Women who stopped the study on March 28 2020 due to COVID 19 pandemic

| | n | tp | fp | tn | fn | Sensitivity % *95% CI* | Specificity % *95% CI* | PPV % *95% CI* | NPV % *95% CI* | PLR *95% CI* | NLR *95% CI* | DOR *95% CI* |
|---|---|---|---|---|---|---|---|---|---|---|---|---|
| HPV | 259 | 50 | 66 | 121 | 22 | 0.69 [0.58, 0.79] | 0.65 [0.58, 0.71] | 0.43 [0.35, 0.52] | 0.84 [0.78, 0.90] | 1.97 [1.53, 2.52] | 0.47 [0.33, 0.68] | 4.17 [2.32, 7.47] |
| VIA | 260 | 16 | 14 | 174 | 56 | 0.22 [0.14, 0.33] | 0.92 [0.88, 0.96] | 0.53 [0.36, 0.70] | 0.76 [0.70, 0.81] | 2.98 [1.54, 5.80] | 0.84 [0.74, 0.96] | 3.55 [1.63, 7.73] |
| Gyncoluar* (youden) | 260 | 35 | 37 | 151 | 37 | 0.48 [0.37, 0.59] | 0.80 [0.74, 0.85] | 0.48 [0.37, 0.60] | 0.80 [0.74, 0.85] | 2.47 [1.70, 3.59] | 0.64 [0.51, 0.81] | 3.86 [2.15, 6.93] |

*AUC: 0.677 [0.603, 0.75] Tp, true positive; fp, false positive; tn, true negative; fn, false negative; PPV, positive predictive value; NPV, negative predictive value; PLR, positive likelihood ratio; NLR, negative likelihood ratio; DOR, diagnostic odds ratio; AUC, area under the receiver operating curve; hrHPV, high-risk human papillomavirus; VIA , visual inspection of the uterine cervix after application of 3–5% acetic, HPV, human papillomavirus test, 95% CI, ninety-five percent confidence interval, n number of women

### C. Missing data

Missing tests excluded (as per main analysis Table 3)

| | n | tp | fp | tn | fn | Sensitivity % *95% CI* | Specificity % *95% CI* | PPV % *95% CI* | NPV % *95% CI* | PLR *95% CI* | NLR *95% CI* | DOR *95% CI* |
|---|---|---|---|---|---|---|---|---|---|---|---|---|
| HPV | 369 | 68 | 93 | 175 | 33 | 0.67 [0.58, 0.76] | 0.65 [0.59, 0.71] | 0.42 [0.35, 0.50] | 0.84 [0.79, 0.88] | 1.94 [1.57, 2.40] | 0.50 [0.37, 0.67] | 3.88 [2.39, 6.30] |
| VIA | 371 | 23 | 20 | 250 | 78 | 0.23 [0.16, 0.32] | 0.93 [0.89, 0.95] | 0.55 [0.39, 0.68] | 0.76 [0.71, 0.81] | 3.07 [1.77, 5.35] | 0.83 [0.75, 0.93] | 3.69 [1.92, 7.07] |
| Gyncoluar* (youden) | 371 | 52 | 54 | 216 | 49 | 0.52 [0.42, 0.61] | 0.80 [0.75, 0.84] | 0.49 [0.40, 0.58] | 0.82 [0.76, 0.86] | 2.57 [1.90, 3.49] | 0.61 [0.49, 0.75] | 4.25 [2.60, 6.94] |

AUC: 0.69 [0.63, 0.75], Tp, true positive; fp, false positive; tn, true negative; fn, false negative; PPV, positive predictive value; NPV, negative predictive value; PLR, positive likelihood ratio; NLR, negative likelihood ratio; DOR, diagnostic odds ratio; AUC, area under the receiver operating curve; hrHPV, high-risk human papillomavirus; VIA , visual inspection of the uterine cervix after application of 3–5% acetic, HPV, human papillomavirus test, 95% CI, ninety-five percent confidence interval, n number of women

Missing tests considered positive

| | n | tp | fp | tn | fn | Sensitivity % *95% CI* | Specificity % *95% CI* | PPV % *95% CI* | NPV % *95% CI* | PLR *95% CI* | NLR *95% CI* | DOR *95% CI* |
|---|---|---|---|---|---|---|---|---|---|---|---|---|
| HPV | 371 | 68 | 95 | 175 | 33 | 0.673 [0.577, 0.757] | 0.648 [0.589, 0.703] | 0.417 [0.344, 0.494] | 0.841 [0.786, 0.885] | 1.913 [1.549, 2.364] | 0.504 [0.376, 0.676] | 3.796 [2.337, 6.166] |
| VIA | 371 | 23 | 20 | 250 | 78 | 0.228 [0.157, 0.319] | 0.926 [0.888, 0.952] | 0.535 [0.389, 0.675] | 0.762 [0.713, 0.805] | 3.074 [1.767, 5.349] | 0.834 [0.746, 0.932] | 3.686 [1.922, 7.067] |
| Gyncoluar* (youden) | 371 | 52 | 54 | 216 | 49 | 0.515 [0.419, 0.61] | 0.8 [0.748, 0.843] | 0.491 [0.397, 0.584] | 0.815 [0.764, 0.857] | 2.574 [1.898, 3.491] | 0.606 [0.492, 0.748] | 4.245 [2.598, 6.937] |

AUC: 0.69 [0.628, 0.751] Tp, true positive; fp, false positive; tn, true negative; fn, false negative; PPV, positive predictive value; NPV, negative predictive value; PLR, positive likelihood ratio; NLR, negative likelihood ratio; DOR, diagnostic odds ratio; AUC, area under the receiver operating curve; hrHPV, high-risk human papillomavirus; VIA , visual inspection of the uterine cervix after application of 3–5% acetic, HPV, human papillomavirus test, 95% CI, ninety-five percent confidence interval, n number of women

Missing tests considered negative

| | n | tp | fp | tn | fn | **Sensitivity %** *95% CI* | **Specificity %** *95% CI* | **PPV %** *95% CI* | **NPV %** *95% CI* | **PLR** *95% CI* | **NLR** *95% CI* | **DOR** *95% CI* |
|---|---|---|---|---|---|---|---|---|---|---|---|---|
| HPV | 371 | 68 | 93 | 177 | 33 | 0.673 [0.577, 0.757] | 0.656 [0.587, 0.710] | 0.422 [0.349, 0.500] | 0.843 [0.788, 0.886] | 1.913 [1.549, 2.364] | 0.498 [0.372, 0.688] | 3.922 [2.413, 6.374] |
| VIA | 371 | 23 | 20 | 250 | 78 | 0.228 [0.157, 0.319] | 0.926 [0.888, 0.952] | 0.535 [0.389, 0.675] | 0.762 [0.713, 0.805] | 3.074 [1.767, 5.349] | 0.834 [0.746, 0.932] | 3.686 [1.922, 7.067] |
| Gyncoluar* (youden) | 371 | 52 | 54 | 216 | 49 | 0.515 [0.419, 0.61] | 0.8 [0.748, 0.843] | 0.491 [0.397, 0.584] | 0.815 [0.764, 0.857] | 2.574 [1.898, 3.491] | 0.606 [0.492, 0.748] | 4.245 [2.598, 6.937] |

*AUC: 0.69 [0.628, 0.751] Tp, true positive; fp, false positive; tn, true negative; fn, false negative; PPV, positive predictive value; NPV, negative predictive value; PLR, positive likelihood ratio; NLR, negative likelihood ratio; DOR, diagnostic odds ratio; AUC, area under the receiver operating curve; hrHPV, high-risk human papillomavirus; VIA , visual inspection of the uterine cervix after application of 3–5% acetic, HPV, human papillomavirus test, 95% CI, ninety-five percent confidence interval, n number of women

### D. Biopsy of visible lesions only

| | n | tp | fp | tn | fn | **Sensitivity %** *95% CI* | **Specificity %** *95% CI* | **PPV %** *95% CI* | **NPV %** *95% CI* | **PLR** *95% CI* | **NLR** *95% CI* | **DOR** *95% CI* |
|---|---|---|---|---|---|---|---|---|---|---|---|---|
| HPV | 371 | 42 | 120 | 202 | 7 | 0.86 [0.73, 0.93] | 0.63 [0.57, 0.68] | 0.30 [0.20, 0.33] | 0.97 [0.93, 0.98] | 2.30 [1.92, 2.76] | 0.23 [0.11, 0.45] | 10.10 [4.40, 23.20] |
| VIA | 373 | 22 | 21 | 303 | 27 | 0.45 [0.32, 0.59] | 0.94 [0.90, 0.96] | 0.51 [0.37, 0.65] | 0.92 [0.88, 0.94] | 6.93 [4.13, 11.62] | 0.60 [0.46, 0.76] | 11.76 [5.75, 24.05] |
| Gyncoluar* (youden) | 373 | 46 | 60 | 264 | 3 | 0.94 [0.84, 0.98] | 0.82 [0.77, 0.85] | 0.43 [0.34, 0.53] | 0.99 [0.97, 0.99] | 5.07 [3.99, 6.44] | 0.08 [0.03, 0.23] | 67.47 [20.30, 224.24] |

*AUC: 0.936 [0.904, 0.968] Tp, true positive; fp, false positive; tn, true negative; fn, false negative; PPV, positive predictive value; NPV, negative predictive value; PLR, positive likelihood ratio; NLR, negative likelihood ratio; DOR, diagnostic odds ratio; AUC, area under the receiver operating curve; hrHPV, high-risk human papillomavirus; VIA , visual inspection of the uterine cervix after application of 3–5% acetic, HPV, human papillomavirus test, 95% CI, ninety-five percent confidence interval, n number of women

Figures

**S12. Supplementary material 12: Sensitivity and specificity of combination test screening strategies for prevalent CIN2**

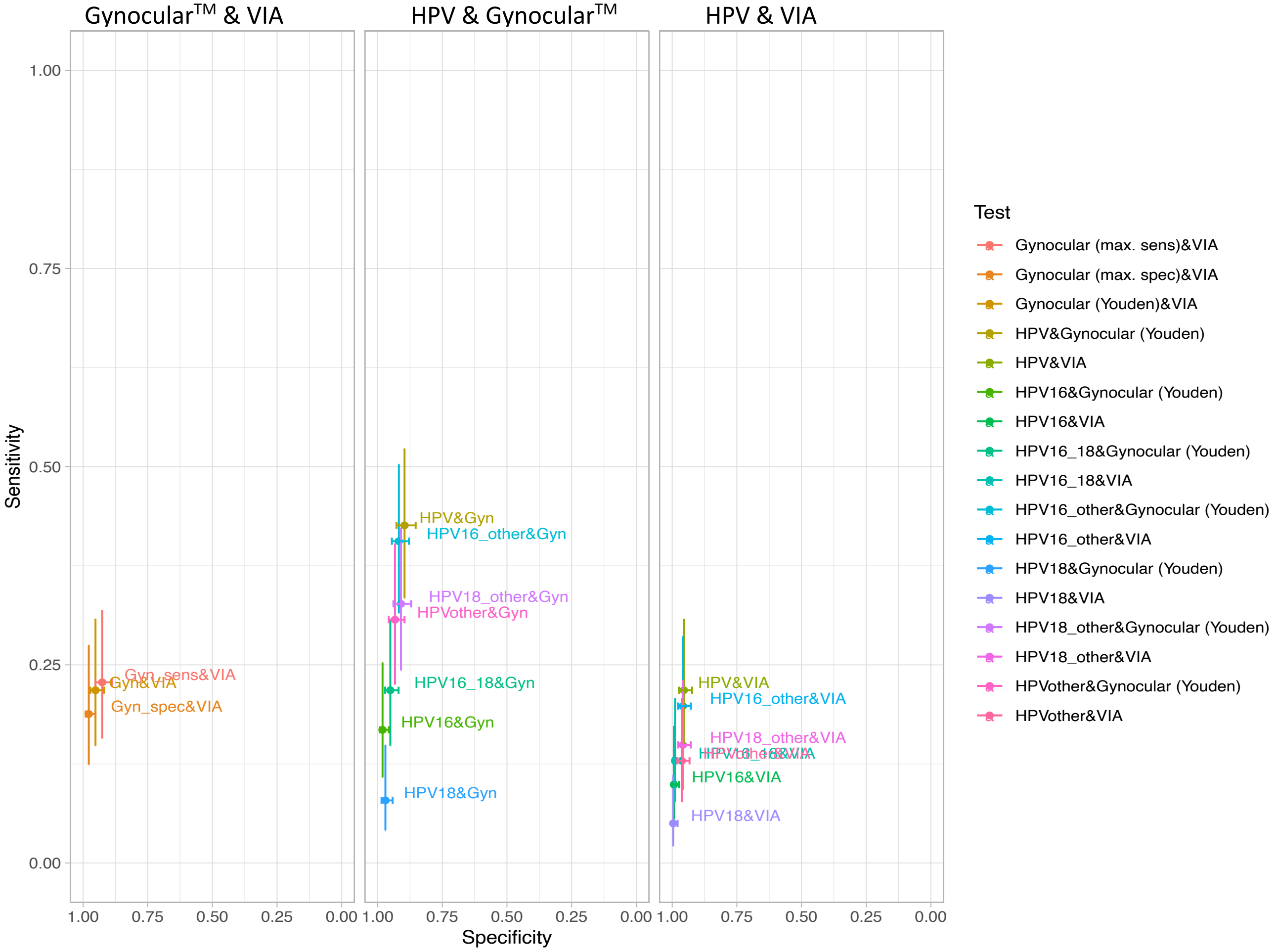

Max.spec= using a threshold that maximises specificity. Max.sens= using a threshold that maximises sensitivity. HPV16= human papillomavirus subtype 16. HPV18= human papillomavirus subtypes 18 and 45. HPVother= human papillomavirus other high-risk subtypes pooled -31, 33, 35, 39, 51, 52, 56, 58, 59, 66, and 68. hrHPV= high-risk human papillomavirus. VIA= visual inspection of the uterine cervix after application of 3–5% acetic.

### S13. Supplementary material 13: Calculation of combined tests – alternate calculation

Only participants who test positive to the first test are included in the denominator in these analyses – this correlates to the clinical scenario when you assess accuracy of a test at a referral site for example a colposcopy clinic.

#### A. Sequential combination of two tests

| t1 | t2 | N | TP | FP | TN | FN | Sens. [CI] | Spec. [CI] | PPV [CI] | NPV [CI] | PLR [CI] | NLR [CI] |
|---|---|---|---|---|---|---|---|---|---|---|---|---|
| HPV (pos.) | Gynocular (Youden) | 161 (neg.:93; pos.:68) | 43 | 28 | 65 | 25 | 0.632 [0.514, 0.737] | 0.699 [0.599, 0.783] | 0.606 [0.489, 0.711] | 0.722 [0.622, 0.804] | 2.1 [1.467, 3.007] | 0.526 [0.375, 0.738] |
| | Gynocular (max. spec) | 161 (neg.:93; pos.:68) | 27 | 10 | 83 | 41 | 0.397 [0.289, 0.516] | 0.892 [0.813, 0.941] | 0.73 [0.57, 0.846] | 0.669 [0.583, 0.746] | 3.693 [1.919, 7.107] | 0.676 [0.55, 0.83] |
| | VIA | 161 (neg.:93; pos.:68) | 22 | 12 | 81 | 46 | 0.324 [0.224, 0.442] | 0.871 [0.788, 0.925] | 0.647 [0.479, 0.785] | 0.638 [0.551, 0.716] | 2.507 [1.335, 4.708] | 0.777 [0.647, 0.932] |
| | HPV16 | 161 (neg.:93; pos.:68) | 20 | 19 | 74 | 48 | 0.294 [0.199, 0.411] | 0.796 [0.703, 0.865] | 0.513 [0.362, 0.661] | 0.607 [0.518, 0.689] | 1.44 [0.835, 2.482] | 0.887 [0.737, 1.067] |
| | HPV16_18 | 161 (neg.:93; pos.:68) | 28 | 40 | 53 | 40 | 0.412 [0.303, 0.53] | 0.57 [0.468, 0.666] | 0.412 [0.303, 0.53] | 0.57 [0.468, 0.666] | 0.957 [0.663, 1.383] | 1.032 [0.791, 1.347] |
| | HPV16_other | 161 (neg.:93; pos.:68) | 65 | 76 | 17 | 3 | 0.956 [0.878, 0.985] | 0.183 [0.117, 0.273] | 0.461 [0.381, 0.543] | 0.85 [0.64, 0.948] | 1.17 [1.049, 1.304] | 0.241 [0.074, 0.791] |
| | HPV18 | 161 (neg.:93; pos.:68) | 11 | 21 | 72 | 57 | 0.162 [0.093, 0.267] | 0.774 [0.679, 0.847] | 0.344 [0.204, 0.517] | 0.558 [0.472, 0.641] | 0.716 [0.371, 1.385] | 1.083 [0.931, 1.26] |
| | HPV18_other | 161 (neg.:93; pos.:68) | 56 | 79 | 14 | 12 | 0.824 [0.716, 0.896] | 0.151 [0.092, 0.237] | 0.415 [0.335, 0.499] | 0.538 [0.355, 0.712] | 0.969 [0.843, 1.114] | 1.172 [0.579, 2.372] |
| | HPVother | 161 (neg.:93; pos.:68) | 53 | 62 | 31 | 15 | 0.779 [0.667, 0.862] | 0.333 [0.246, 0.434] | 0.461 [0.373, 0.552] | 0.674 [0.53, 0.791] | 1.169 [0.965, 1.416] | 0.662 [0.389, 1.126] |

| t1 | t2 | N | TP | FP | TN | FN | Sens. [CI] | Spec. [CI] | PPV [CI] | NPV [CI] | PLR [CI] | NLR [CI] |
|---|---|---|---|---|---|---|---|---|---|---|---|---|
| HPV16 (pos.) | Gynocular (Youden) | 39 (neg.:19; pos.:20) | 17 | 5 | 14 | 3 | 0.85 [0.64, 0.948] | 0.737 [0.512, 0.882] | 0.773 [0.566, 0.899] | 0.824 [0.59, 0.938] | 3.23 [1.489, 7.008] | 0.204 [0.069, 0.598] |
| | Gynocular (max. spec) | 39 (neg.:19; pos.:20) | 11 | 2 | 17 | 9 | 0.55 [0.342, 0.742] | 0.895 [0.686, 0.971] | 0.846 [0.578, 0.957] | 0.654 [0.462, 0.806] | 5.225 [1.328, 20.553] | 0.503 [0.302, 0.836] |
| | VIA | 39 (neg.:19; pos.:20) | 10 | 2 | 17 | 10 | 0.5 [0.299, 0.701] | 0.895 [0.686, 0.971] | 0.833 [0.552, 0.953] | 0.63 [0.442, 0.785] | 4.75 [1.192, 18.923] | 0.559 [0.351, 0.889] |
| HPV16_18 (pos.) | Gynocular (Youden) | 68 (neg.:40; pos.:28) | 22 | 13 | 27 | 6 | 0.786 [0.605, 0.898] | 0.675 [0.52, 0.799] | 0.629 [0.463, 0.768] | 0.818 [0.656, 0.914] | 2.418 [1.486, 3.933] | 0.317 [0.151, 0.666] |
| | Gynocular (max. spec) | 68 (neg.:40; pos.:28) | 14 | 4 | 36 | 14 | 0.5 [0.326, 0.674] | 0.9 [0.769, 0.96] | 0.778 [0.548, 0.91] | 0.72 [0.583, 0.825] | 5 [1.838, 13.602] | 0.556 [0.378, 0.816] |
| | VIA | 68 (neg.:40; pos.:28) | 13 | 3 | 37 | 15 | 0.464 [0.026, 0.199] | 0.925 [0.358, 0.705] | 0.812 [0.066, 0.43] | 0.712 [0.183, 0.423] | 6.19 [0.051, 0.515] | 0.579 [1.21, 2.465] |
| HPV16_other (pos.) | Gynocular (Youden) | 141 (neg.:76; pos.:65) | 41 | 22 | 54 | 24 | 0.631 [0.509, 0.738] | 0.711 [0.6, 0.8] | 0.651 [0.528, 0.757] | 0.692 [0.583, 0.784] | 2.179 [1.463, 3.245] | 0.52 [0.367, 0.736] |
| | Gynocular (max. spec) | 141 (neg.:76; pos.:65) | 25 | 8 | 68 | 40 | 0.385 [0.276, 0.506] | 0.895 [0.806, 0.946] | 0.758 [0.59, 0.872] | 0.63 [0.536, 0.715] | 3.654 [1.771, 7.537] | 0.688 [0.559, 0.846] |
| | VIA | 141 (neg.:76; pos.:65) | 20 | 11 | 65 | 45 | 0.308 [0.209, 0.428] | 0.855 [0.759, 0.917] | 0.645 [0.469, 0.789] | 0.591 [0.497, 0.678] | 2.126 [1.102, 4.101] | 0.809 [0.672, 0.976] |
| HPV18 (pos.) | Gynocular (Youden) | 32 (neg.:21; pos.:11) | 8 | 8 | 13 | 3 | 0.727 [0.434, 0.903] | 0.619 [0.409, 0.792] | 0.5 [0.28, 0.72] | 0.812 [0.57, 0.934] | 1.909 [0.992, 3.673] | 0.441 [0.159, 1.224] |
| | Gynocular (max. spec) | 32 (neg.:21; pos.:11) | 5 | 2 | 19 | 6 | 0.455 [0.027, 0.289] | 0.905 [0.28, 0.787] | 0.714 [0.082, 0.641] | 0.76 [0.115, 0.434] | 4.773 [0.048, 0.91] | 0.603 [0.95, 2.895] |

| t1 | t2 | N | TP | FP | TN | FN | Sens. [CI] | Spec. [CI] | PPV [CI] | NPV [CI] | PLR [CI] | NLR [CI] |
|---|---|---|---|---|---|---|---|---|---|---|---|---|
| | VIA | 32 (neg.:21; pos.:11) | 5 | 1 | 20 | 6 | 0.455 [0.008, 0.227] | 0.952 [0.28, 0.787] | 0.833 [0.03, 0.564] | 0.769 [0.11, 0.421] | 9.545 [0.014, 0.789] | 0.573 [1.01, 3.02] |
| HPV18_other (pos.) | Gynocular (Youden) | 135 (neg.:79; pos.:56) | 33 | 24 | 55 | 23 | 0.589 [0.459, 0.708] | 0.696 [0.588, 0.787] | 0.579 [0.45, 0.698] | 0.705 [0.596, 0.795] | 1.94 [1.301, 2.891] | 0.59 [0.417, 0.834] |
| | Gynocular (max. spec) | 135 (neg.:79; pos.:56) | 20 | 9 | 70 | 36 | 0.357 [0.245, 0.488] | 0.886 [0.797, 0.939] | 0.69 [0.508, 0.827] | 0.66 [0.566, 0.744] | 3.135 [1.544, 6.366] | 0.726 [0.588, 0.896] |
| | VIA | 135 (neg.:79; pos.:56) | 15 | 11 | 68 | 41 | 0.268 [0.17, 0.396] | 0.861 [0.768, 0.92] | 0.577 [0.389, 0.745] | 0.624 [0.53, 0.709] | 1.924 [0.957, 3.869] | 0.851 [0.709, 1.02] |
| HPVother (pos.) | Gynocular (Youden) | 115 (neg.:62; pos.:53) | 31 | 18 | 44 | 22 | 0.585 [0.451, 0.707] | 0.71 [0.587, 0.808] | 0.633 [0.493, 0.753] | 0.667 [0.547, 0.768] | 2.015 [1.284, 3.161] | 0.585 [0.409, 0.836] |
| | Gynocular (max. spec) | 115 (neg.:62; pos.:53) | 18 | 7 | 55 | 35 | 0.34 [0.227, 0.474] | 0.887 [0.785, 0.944] | 0.72 [0.524, 0.857] | 0.611 [0.508, 0.705] | 3.008 [1.362, 6.643] | 0.744 [0.602, 0.921] |
| | VIA | 115 (neg.:62; pos.:53) | 13 | 10 | 52 | 40 | 0.245 [0.149, 0.376] | 0.839 [0.728, 0.91] | 0.565 [0.368, 0.744] | 0.565 [0.463, 0.662] | 1.521 [0.727, 3.182] | 0.9 [0.745, 1.086] |
| Gynocular (max. sens) (pos.) | HPV | 358 (neg.:260; pos.:98) | 67 | 91 | 169 | 31 | 0.684 [0.586, 0.767] | 0.65 [0.59, 0.705] | 0.424 [0.35, 0.502] | 0.845 [0.788, 0.889] | 1.953 [1.578, 2.418] | 0.487 [0.359, 0.66] |
| | HPV16 | 358 (neg.:260; pos.:98) | 20 | 18 | 242 | 78 | 0.204 [0.136, 0.294] | 0.931 [0.893, 0.956] | 0.526 [0.373, 0.675] | 0.756 [0.706, 0.8] | 2.948 [1.629, 5.333] | 0.855 [0.769, 0.95] |
| | HPV16_18 | 358 (neg.:260; pos.:98) | 28 | 39 | 221 | 70 | 0.286 [0.206, 0.382] | 0.85 [0.802, 0.888] | 0.418 [0.307, 0.537] | 0.759 [0.707, 0.805] | 1.905 [1.244, 2.917] | 0.84 [0.734, 0.962] |
| | HPV16_other | 358 (neg.:260; pos.:98) | 64 | 74 | 186 | 34 | 0.653 [0.555, 0.74] | 0.715 [0.658, 0.767] | 0.464 [0.383, 0.547] | 0.845 [0.792, 0.887] | 2.295 [1.804, 2.919] | 0.485 [0.366, 0.643] |

| t1 | t2 | N | TP | FP | TN | FN | Sens. [CI] | Spec. [CI] | PPV [CI] | NPV [CI] | PLR [CI] | NLR [CI] |
|---|---|---|---|---|---|---|---|---|---|---|---|---|
| | HPV18 | 358 (neg.:260; pos.:98) | 11 | 21 | 239 | 87 | 0.112 [0.064, 0.19] | 0.919 [0.88, 0.947] | 0.344 [0.204, 0.517] | 0.733 [0.683, 0.778] | 1.39 [0.696, 2.775] | 0.966 [0.892, 1.045] |
| | HPV18_other | 358 (neg.:260; pos.:98) | 55 | 78 | 182 | 43 | 0.561 [0.463, 0.655] | 0.7 [0.642, 0.752] | 0.414 [0.333, 0.499] | 0.809 [0.752, 0.855] | 1.871 [1.449, 2.415] | 0.627 [0.494, 0.795] |
| | HPVother | 358 (neg.:260; pos.:98) | 52 | 61 | 199 | 46 | 0.531 [0.433, 0.626] | 0.765 [0.71, 0.813] | 0.46 [0.371, 0.552] | 0.812 [0.759, 0.856] | 2.262 [1.696, 3.016] | 0.613 [0.492, 0.765] |
| Gynocular (max. spec) (neg.) | HPV | 323 (neg.:252; pos.:71) | 41 | 83 | 169 | 30 | 0.577 [0.462, 0.685] | 0.671 [0.61, 0.726] | 0.331 [0.254, 0.417] | 0.849 [0.793, 0.892] | 1.753 [1.344, 2.287] | 0.63 [0.474, 0.838] |
| | HPV16 | 323 (neg.:252; pos.:71) | 9 | 17 | 235 | 62 | 0.127 [0.068, 0.224] | 0.933 [0.895, 0.957] | 0.346 [0.194, 0.538] | 0.791 [0.741, 0.834] | 1.879 [0.875, 4.033] | 0.936 [0.852, 1.029] |
| | HPV16_18 | 323 (neg.:252; pos.:71) | 14 | 36 | 216 | 57 | 0.197 [0.121, 0.304] | 0.857 [0.809, 0.895] | 0.28 [0.175, 0.417] | 0.791 [0.739, 0.835] | 1.38 [0.79, 2.412] | 0.937 [0.826, 1.062] |
| | HPV16_other | 323 (neg.:252; pos.:71) | 40 | 68 | 184 | 31 | 0.563 [0.448, 0.673] | 0.73 [0.672, 0.781] | 0.37 [0.285, 0.464] | 0.856 [0.803, 0.897] | 2.088 [1.565, 2.786] | 0.598 [0.454, 0.787] |
| | HPV18 | 323 (neg.:252; pos.:71) | 6 | 19 | 233 | 65 | 0.085 [0.039, 0.172] | 0.925 [0.885, 0.951] | 0.24 [0.115, 0.434] | 0.782 [0.732, 0.825] | 1.121 [0.465, 2.7] | 0.99 [0.915, 1.072] |
| | HPV18_other | 323 (neg.:252; pos.:71) | 36 | 70 | 182 | 35 | 0.507 [0.393, 0.62] | 0.722 [0.664, 0.774] | 0.34 [0.256, 0.434] | 0.839 [0.784, 0.882] | 1.825 [1.347, 2.473] | 0.683 [0.533, 0.875] |
| | HPVother | 323 (neg.:252; pos.:71) | 35 | 55 | 197 | 36 | 0.493 [0.38, 0.607] | 0.782 [0.727, 0.828] | 0.389 [0.295, 0.492] | 0.845 [0.794, 0.886] | 2.259 [1.62, 3.148] | 0.649 [0.511, 0.823] |
| Gynocular (Youden) (neg.) | HPV | 263 (neg.:214; pos.:49) | 25 | 65 | 149 | 24 | 0.51 [0.375, 0.644] | 0.696 [0.632, 0.754] | 0.278 [0.196, 0.378] | 0.861 [0.802, 0.905] | 1.68 [1.194, 2.363] | 0.703 [0.522, 0.949] |

| t1 | t2 | N | TP | FP | TN | FN | Sens. [CI] | Spec. [CI] | PPV [CI] | NPV [CI] | PLR [CI] | NLR [CI] |
|---|---|---|---|---|---|---|---|---|---|---|---|---|
| | HPV16 | 263 (neg.:214; pos.:49) | 3 | 14 | 200 | 46 | 0.061 [0.021, 0.165] | 0.935 [0.893, 0.961] | 0.176 [0.062, 0.41] | 0.813 [0.76, 0.857] | 0.936 [0.28, 3.131] | 1.004 [0.927, 1.088] |
| | HPV16_18 | 263 (neg.:214; pos.:49) | 6 | 27 | 187 | 43 | 0.122 [0.057, 0.242] | 0.874 [0.823, 0.912] | 0.182 [0.086, 0.344] | 0.813 [0.758, 0.858] | 0.971 [0.424, 2.222] | 1.004 [0.894, 1.128] |
| | HPV16_othe r | 263 (neg.:214; pos.:49) | 24 | 54 | 160 | 25 | 0.49 [0.356, 0.625] | 0.748 [0.685, 0.801] | 0.308 [0.216, 0.417] | 0.865 [0.808, 0.907] | 1.941 [1.344, 2.802] | 0.682 [0.513, 0.908] |
| | HPV18 | 263 (neg.:214; pos.:49) | 3 | 13 | 201 | 46 | 0.061 [0.021, 0.165] | 0.939 [0.899, 0.964] | 0.188 [0.066, 0.43] | 0.814 [0.761, 0.857] | 1.008 [0.299, 3.402] | 0.999 [0.923, 1.082] |
| | HPV18 othe r | 263 (neg.:214; pos.:49) | 23 | 55 | 159 | 26 | 0.469 [0.337, 0.606] | 0.743 [0.681, 0.797] | 0.295 [0.205, 0.404] | 0.859 [0.802, 0.902] | 1.826 [1.255, 2.657] | 0.714 [0.543, 0.94] |
| | HPVother | 263 (neg.:214; pos.:49) | 22 | 44 | 170 | 27 | 0.449 [0.319, 0.587] | 0.794 [0.735, 0.843] | 0.333 [0.232, 0.453] | 0.863 [0.808, 0.904] | 2.184 [1.454, 3.28] | 0.694 [0.534, 0.901] |
| | HPV | 106 (neg.:54; pos.:52) | 43 | 28 | 26 | 9 | 0.827 [0.703, 0.906] | 0.481 [0.354, 0.611] | 0.606 [0.489, 0.711] | 0.743 [0.579, 0.858] | 1.595 [1.199, 2.122] | 0.359 [0.187, 0.692] |
| | HPV16 | 106 (neg.:54; pos.:52) | 17 | 5 | 49 | 35 | 0.327 [0.215, 0.462] | 0.907 [0.801, 0.96] | 0.773 [0.566, 0.899] | 0.583 [0.477, 0.683] | 3.531 [1.405, 8.873] | 0.742 [0.603, 0.913] |
| Gynocular (Youden) (pos.) | HPV16_18 | 106 (neg.:54; pos.:52) | 22 | 13 | 41 | 30 | 0.423 [0.299, 0.558] | 0.759 [0.631, 0.854] | 0.629 [0.463, 0.768] | 0.577 [0.462, 0.685] | 1.757 [0.994, 3.108] | 0.76 [0.576, 1.002] |
| | HPV16_othe r | 106 (neg.:54; pos.:52) | 41 | 22 | 32 | 11 | 0.788 [0.66, 0.878] | 0.593 [0.46, 0.713] | 0.651 [0.528, 0.757] | 0.744 [0.598, 0.851] | 1.935 [1.362, 2.749] | 0.357 [0.202, 0.631] |
| | HPV18 | 106 (neg.:54; pos.:52) | 8 | 8 | 46 | 44 | 0.154 [0.08, 0.275] | 0.852 [0.734, 0.923] | 0.5 [0.28, 0.72] | 0.511 [0.41, 0.612] | 1.038 [0.421, 2.562] | 0.993 [0.846, 1.166] |

| t1 | t2 | N | TP | FP | TN | FN | Sens. [CI] | Spec. [CI] | PPV [CI] | NPV [CI] | PLR [CI] | NLR [CI] |
|---|---|---|---|---|---|---|---|---|---|---|---|---|
| | HPV18_other | 106 (neg.:54; pos.:52) | 33 | 24 | 30 | 19 | 0.635 [0.499, 0.752] | 0.556 [0.424, 0.68] | 0.579 [0.45, 0.698] | 0.612 [0.472, 0.736] | 1.428 [0.994, 2.052] | 0.658 [0.428, 1.011] |
| | HPVother | 106 (neg.:54; pos.:52) | 31 | 18 | 36 | 21 | 0.596 [0.461, 0.718] | 0.667 [0.534, 0.778] | 0.633 [0.493, 0.753] | 0.632 [0.502, 0.745] | 1.788 [1.154, 2.773] | 0.606 [0.414, 0.886] |
| VIA (neg.) | Gynocular (Youden) | 328 (neg.:250; pos.:78) | 30 | 41 | 209 | 48 | 0.385 [0.284, 0.496] | 0.836 [0.785, 0.877] | 0.423 [0.315, 0.538] | 0.813 [0.761, 0.856] | 2.345 [1.578, 3.486] | 0.736 [0.612, 0.885] |
| | Gynocular (max. spec) | 328 (neg.:250; pos.:78) | 11 | 10 | 240 | 67 | 0.141 [0.081, 0.235] | 0.96 [0.928, 0.978] | 0.524 [0.324, 0.717] | 0.782 [0.732, 0.824] | 3.526 [1.556, 7.987] | 0.895 [0.815, 0.982] |
| | HPV | 326 (neg.:248; pos.:78) | 46 | 81 | 167 | 32 | 0.59 [0.479, 0.692] | 0.673 [0.613, 0.729] | 0.362 [0.284, 0.449] | 0.839 [0.782, 0.884] | 1.806 [1.396, 2.335] | 0.609 [0.461, 0.806] |
| | HPV16 | 326 (neg.:248; pos.:78) | 10 | 17 | 231 | 68 | 0.128 [0.071, 0.22] | 0.931 [0.893, 0.957] | 0.37 [0.215, 0.558] | 0.773 [0.722, 0.816] | 1.87 [0.894, 3.914] | 0.936 [0.854, 1.026] |
| | HPV16_18 | 326 (neg.:248; pos.:78) | 15 | 37 | 211 | 63 | 0.192 [0.12, 0.293] | 0.851 [0.801, 0.89] | 0.288 [0.183, 0.423] | 0.77 [0.717, 0.816] | 1.289 [0.749, 2.219] | 0.949 [0.842, 1.071] |
| | HPV16_other | 326 (neg.:248; pos.:78) | 45 | 65 | 183 | 33 | 0.577 [0.466, 0.68] | 0.738 [0.68, 0.789] | 0.409 [0.322, 0.503] | 0.847 [0.793, 0.889] | 2.201 [1.66, 2.919] | 0.573 [0.438, 0.751] |
| | HPV18 | 326 (neg.:248; pos.:78) | 6 | 20 | 228 | 72 | 0.077 [0.036, 0.158] | 0.919 [0.879, 0.947] | 0.231 [0.11, 0.421] | 0.76 [0.709, 0.805] | 0.954 [0.397, 2.291] | 1.004 [0.933, 1.081] |
| | HPV18_other | 326 (neg.:248; pos.:78) | 41 | 68 | 180 | 37 | 0.526 [0.416, 0.633] | 0.726 [0.667, 0.778] | 0.376 [0.291, 0.47] | 0.829 [0.774, 0.874] | 1.917 [1.431, 2.568] | 0.654 [0.511, 0.836] |
| | HPVother | 326 (neg.:248; pos.:78) | 40 | 52 | 196 | 38 | 0.513 [0.404, 0.621] | 0.79 [0.735, 0.836] | 0.435 [0.338, 0.537] | 0.838 [0.785, 0.879] | 2.446 [1.768, 3.383] | 0.616 [0.487, 0.781] |

| t1 | t2 | N | TP | FP | TN | FN | Sens. [CI] | Spec. [CI] | PPV [CI] | NPV [CI] | PLR [CI] | NLR [CI] |
|---|---|---|---|---|---|---|---|---|---|---|---|---|
| VIA (pos.) | Gynocular (Youden) | 43 (neg.:20; pos.:23) | 22 | 13 | 7 | 1 | 0.957 [0.79, 0.992] | 0.35 [0.181, 0.567] | 0.629 [0.463, 0.768] | 0.875 [0.529, 0.978] | 1.472 [1.055, 2.053] | 0.124 [0.017, 0.925] |
| | Gynocular (max. spec) | 43 (neg.:20; pos.:23) | 19 | 6 | 14 | 4 | 0.826 [0.629, 0.93] | 0.7 [0.481, 0.855] | 0.76 [0.566, 0.885] | 0.778 [0.548, 0.91] | 2.754 [1.374, 5.519] | 0.248 [0.097, 0.633] |
| | HPV | 43 (neg.:20; pos.:23) | 22 | 12 | 8 | 1 | 0.957 [0.79, 0.992] | 0.4 [0.219, 0.613] | 0.647 [0.479, 0.785] | 0.889 [0.565, 0.98] | 1.594 [1.103, 2.304] | 0.109 [0.015, 0.796] |
| | HPV16 | 43 (neg.:20; pos.:23) | 10 | 2 | 18 | 13 | 0.435 [0.256, 0.632] | 0.9 [0.699, 0.972] | 0.833 [0.552, 0.953] | 0.581 [0.408, 0.736] | 4.348 [1.078, 17.542] | 0.628 [0.426, 0.925] |
| | HPV16_18 | 43 (neg.:20; pos.:23) | 13 | 3 | 17 | 10 | 0.565 [0.368, 0.744] | 0.85 [0.64, 0.948] | 0.812 [0.57, 0.934] | 0.63 [0.442, 0.785] | 3.768 [1.25, 11.356] | 0.512 [0.31, 0.844] |
| | HPV16_other | 43 (neg.:20; pos.:23) | 20 | 11 | 9 | 3 | 0.87 [0.679, 0.955] | 0.45 [0.258, 0.658] | 0.645 [0.469, 0.789] | 0.75 [0.468, 0.911] | 1.581 [1.032, 2.423] | 0.29 [0.091, 0.926] |
| | HPV18 | 43 (neg.:20; pos.:23) | 5 | 1 | 19 | 18 | 0.217 [0.097, 0.419] | 0.95 [0.764, 0.991] | 0.833 [0.436, 0.97] | 0.514 [0.359, 0.666] | 4.348 [0.553, 34.171] | 0.824 [0.65, 1.045] |
| | HPV18_other | 43 (neg.:20; pos.:23) | 15 | 11 | 9 | 8 | 0.652 [0.449, 0.812] | 0.45 [0.258, 0.658] | 0.577 [0.389, 0.745] | 0.529 [0.31, 0.738] | 1.186 [0.722, 1.948] | 0.773 [0.369, 1.62] |
| | HPVother | 43 (neg.:20; pos.:23) | 13 | 10 | 10 | 10 | 0.565 [0.368, 0.744] | 0.5 [0.299, 0.701] | 0.565 [0.368, 0.744] | 0.5 [0.299, 0.701] | 1.13 [0.642, 1.991] | 0.87 [0.459, 1.649] |

### S14. Supplementary material 14: Subgroup analyses

#### A. HPV test

| Primary measures | | | | | |
|---|---|---|---|---|---|
| **Data** | **group** | **N** | **Sens [CI]** | **Spec [CI]** | **ratio-sens** |
| Age | [0,25] | 33 (0:22; 1:11) | | | |
| | (25,35] | 122 (0:87; 1:35) | 0.686 [0.52, 0.814] | 0.575 [0.47, 0.673] | ref |
| | (35,45] | 133 (0:98; 1:35) | 0.657 [0.492, 0.792] | 0.724 [0.629, 0.803] | 0.96 [0.69, 1.33] |
| | (45,70] | 75 (0:57; 1:18) | 0.667 [0.437, 0.837] | 0.719 [0.592, 0.819] | 0.97 [0.65, 1.44] |
| Menopause | No | 317 (0:227; 1:90) | 0.689 [0.587, 0.775] | 0.648 [0.583, 0.707] | ref |
| | Yes | 52 (0:41; 1:11) | 0.545 [0.28, 0.787] | 0.683 [0.53, 0.804] | 0.79 [0.45, 1.38] |
| Education | Did not finished secondary | 303 (0:217; 1:86) | 0.674 [0.57, 0.764] | 0.664 [0.598, 0.723] | ref |
| | Finished secondary | 53 (0:41; 1:12) | 0.667 [0.391, 0.862] | 0.585 [0.434, 0.722] | 0.99 [0.65, 1.51] |
| | More than secondary | 13 (0:10; 1:3) | | | |
| Contraception method | Condoms method | 36 (0:30; 1:6) | | | |
| | long acting reversible contraception | 86 (0:61; 1:25) | 0.64 [0.445, 0.798] | 0.689 [0.564, 0.791] | ref |
| | Oral hormonal | 16 (0:13; 1:3) | | | |
| | Withdrawal & others | 4 (0:3; 1:1) | | | |
| | none | 227 (0:161; 1:66) | 0.682 [0.562, 0.782] | 0.64 [0.563, 0.71] | 1.07 [0.76, 1.49] |
| Parity categories | 0 | 11 (0:8; 1:3) | | | |

| | | | | | |
|---|---|---|---|---|---|
| | 1-3 | 181 (0:122; 1:59) | 0.746 [0.622, 0.839] | 0.615 [0.526, 0.696] | ref |
| | >3 | 177 (0:138; 1:39) | 0.59 [0.434, 0.729] | 0.674 [0.592, 0.746] | 0.79 [0.59, 1.07] |
| Trichomoniasis result | Negative | 300 (0:221; 1:79) | 0.671 [0.561, 0.764] | 0.656 [0.591, 0.716] | ref |
| | Positive | 69 (0:47; 1:22) | 0.682 [0.473, 0.836] | 0.638 [0.495, 0.76] | 1.02 [0.73, 1.41] |
| | Undetermined | 0 (:) | | | |
| On ART | No | 1 (0:1) | | | |
| | Yes | 368 (0:267; 1:101) | 0.673 [0.577, 0.757] | 0.652 [0.593, 0.706] | ref |
| CD4 count [cells/mm3] | <200 | 12 (0:5; 1:7) | | | |
| | [200,350) | 44 (0:32; 1:12) | 0.75 [0.468, 0.911] | 0.562 [0.393, 0.718] | ref |
| | [350,500) | 95 (0:70; 1:25) | 0.6 [0.407, 0.766] | 0.629 [0.511, 0.732] | 0.8 [0.51, 1.26] |
| | 500 and more | 217 (0:160; 1:57) | 0.649 [0.519, 0.76] | 0.675 [0.599, 0.743] | 0.87 [0.59, 1.26] |
| HIV RNA load [copies/mL] | < 1000 copies/ml | 340 (0:251; 1:89) | 0.64 [0.537, 0.732] | 0.665 [0.605, 0.721] | ref |
| | 1000 and more | 29 (0:17; 1:12) | | | |
| History of treatment for precancer | No | 362 (0:263; 1:99) | 0.667 [0.569, 0.752] | 0.65 [0.591, 0.705] | ref |
| | Yes | 7 (0:5; 1:2) | | | |

B. **VIA**

| **Primary measures** |
|---|

| Data | group | N | Sens [CI] | Spec [CI] | ratio-sens |
|---|---|---|---|---|---|
| Age | [0,25] | 33 (0:22; 1:11) | | | |
| | (25,35] | 123 (0:88; 1:35) | 0.286 [0.163, 0.451] | 0.932 [0.859, 0.968] | ref |
| | (35,45] | 133 (0:98; 1:35) | 0.2 [0.1, 0.359] | 0.949 [0.886, 0.978] | 0.7 [0.3, 1.63] |
| | (45,70] | 75 (0:57; 1:18) | 0.333 [0.163, 0.563] | 0.93 [0.833, 0.972] | 1.17 [0.5, 2.7] |
| Menopause | No | 319 (0:229; 1:90) | 0.244 [0.167, 0.342] | 0.921 [0.879, 0.95] | ref |
| | Yes | 52 (0:41; 1:11) | 0.091 [0.016, 0.377] | 0.951 [0.839, 0.987] | 0.37 [0.06, 2.5] |
| Education | Did not finished secondary | 305 (0:219; 1:86) | 0.209 [0.137, 0.307] | 0.918 [0.874, 0.947] | ref |
| | Finished secondary | 53 (0:41; 1:12) | 0.417 [0.193, 0.68] | 0.951 [0.839, 0.987] | 1.99 [0.91, 4.37] |
| | More than secondary | 13 (0:10; 1:3) | | | |
| Contraception method | Condoms method | 36 (0:30; 1:6) | | | |
| | long-acting reversible contraception | 88 (0:63; 1:25) | 0.12 [0.042, 0.3] | 0.952 [0.869, 0.984] | ref |
| | Oral hormonal | 16 (0:13; 1:3) | | | |
| | Withdrawal & others | 4 (0:3; 1:1) | | | |
| | none | 227 (0:161; 1:66) | 0.273 [0.18, 0.39] | 0.907 [0.852, 0.943] | 2.27 [0.73, 7.05] |
| Parity categories | 0 | 11 (0:8; 1:3) | | | |
| | 1-3 | 183 (0:124; 1:59) | 0.254 [0.161, 0.378] | 0.911 [0.848, 0.95] | ref |

| | | | | | |
|---|---|---|---|---|---|
| | >3 | 177 (0:138; 1:39) | 0.205 [0.108, 0.355] | 0.942 [0.89, 0.97] | 0.81 [0.38, 1.72] |
| Trichomoniasis result | Negative | 302 (0:223; 1:79) | 0.228 [0.149, 0.332] | 0.919 [0.876, 0.948] | ref |
| | Positive | 69 (0:47; 1:22) | 0.227 [0.101, 0.434] | 0.957 [0.858, 0.988] | 1 [0.42, 2.38] |
| | Undetermined | 0 (:) | | | |
| On ART | No | 1 (0:1) | | | |
| | Yes | 370 (0:269; 1:101) | 0.228 [0.157, 0.319] | 0.926 [0.888, 0.951] | ref |
| CD4 count [cells/mm3] | <200 | 12 (0:5; 1:7) | | | |
| | [200,350) | 44 (0:32; 1:12) | 0.25 [0.089, 0.532] | 0.844 [0.682, 0.931] | ref |
| | [350,500) | 95 (0:70; 1:25) | 0.16 [0.064, 0.347] | 0.943 [0.862, 0.978] | 0.64 [0.17, 2.42] |
| | 500 and more | 219 (0:162; 1:57) | 0.193 [0.111, 0.313] | 0.932 [0.883, 0.962] | 0.77 [0.25, 2.35] |
| HIV RNA load [copies/mL] | < 1000 copies/ml | 342 (0:253; 1:89) | 0.213 [0.141, 0.31] | 0.933 [0.895, 0.958] | ref |
| | 1000 and more | 29 (0:17; 1:12) | | | |
| History of treatment for precancer | No | 364 (0:265; 1:99) | 0.222 [0.152, 0.314] | 0.925 [0.886, 0.951] | ref |
| | Yes | 7 (0:5; 1:2) | | | |

C. **Gynocular**

| **Primary measures** | | | | | |
|---|---|---|---|---|---|
| **Data** | **group** | **N** | **Sens [CI]** | **Spec [CI]** | **ratio-sens** |

| | | | | | |
|---|---|---|---|---|---|
| Age | [0,25] | 33 (0:22; 1:11) | | | |
| | (25,35] | 123 (0:88; 1:35) | 0.514 [0.356, 0.67] | 0.784 [0.687, 0.857] | ref |
| | (35,45] | 133 (0:98; 1:35) | 0.486 [0.33, 0.644] | 0.786 [0.695, 0.855] | 0.94 [0.59, 1.51] |
| | (45,70] | 75 (0:57; 1:18) | 0.611 [0.386, 0.797] | 0.86 [0.747, 0.927] | 1.19 [0.73, 1.94] |
| Menopause | No | 319 (0:229; 1:90) | 0.522 [0.42, 0.622] | 0.799 [0.742, 0.846] | ref |
| | Yes | 52 (0:41; 1:11) | 0.455 [0.213, 0.72] | 0.805 [0.66, 0.898] | 0.87 [0.44, 1.71] |
| Education | Did not finished secondary | 305 (0:219; 1:86) | 0.535 [0.43, 0.637] | 0.813 [0.756, 0.859] | ref |
| | Finished secondary | 53 (0:41; 1:12) | 0.5 [0.254, 0.746] | 0.732 [0.581, 0.843] | 0.93 [0.51, 1.7] |
| | More than secondary | 13 (0:10; 1:3) | | | |
| Contraception method | Condoms method | 36 (0:30; 1:6) | | | |
| | long acting reversible contraception | 88 (0:63; 1:25) | 0.44 [0.267, 0.629] | 0.841 [0.732, 0.911] | ref |
| | Oral hormonal | 16 (0:13; 1:3) | | | |
| | Withdrawal & others | 4 (0:3; 1:1) | | | |
| | none | 227 (0:161; 1:66) | 0.5 [0.383, 0.617] | 0.776 [0.706, 0.834] | 1.14 [0.69, 1.88] |
| Parity categories | 0 | 11 (0:8; 1:3) | | | |
| | 1-3 | 183 (0:124; 1:59) | 0.508 [0.384, 0.632] | 0.79 [0.71, 0.853] | ref |
| | >3 | 177 (0:138; 1:39) | 0.564 [0.41, 0.707] | 0.812 [0.738, 0.868] | 1.11 [0.76, 1.61] |

| | | | | | |
|---|---|---|---|---|---|
| Trichomoniasis result | Negative | 302 (0:223; 1:79) | 0.506 [0.398, 0.614] | 0.807 [0.75, 0.854] | ref |
| | Positive | 69 (0:47; 1:22) | 0.545 [0.347, 0.731] | 0.766 [0.628, 0.864] | 1.08 [0.69, 1.67] |
| | Undetermined | 0 (:) | | | |
| On ART | No | 1 (0:1) | | | |
| | Yes | 370 (0:269; 1:101) | 0.515 [0.419, 0.61] | 0.799 [0.747, 0.843] | ref |
| CD4 count [cells/mm3] | <200 | 12 (0:5; 1:7) | | | |
| | [200,350) | 44 (0:32; 1:12) | 0.667 [0.391, 0.862] | 0.75 [0.579, 0.867] | ref |
| | [350,500) | 95 (0:70; 1:25) | 0.4 [0.234, 0.593] | 0.757 [0.645, 0.842] | 0.6 [0.32, 1.12] |
| | 500 and more | 219 (0:162; 1:57) | 0.491 [0.366, 0.617] | 0.827 [0.762, 0.878] | 0.74 [0.46, 1.19] |
| HIV RNA load [copies/mL] | < 1000 copies/ml | 342 (0:253; 1:89) | 0.494 [0.393, 0.596] | 0.822 [0.77, 0.864] | ref |
| | 1000 and more | 29 (0:17; 1:12) | | | |
| History of treatment for precancer | No | 364 (0:265; 1:99) | 0.505 [0.408, 0.601] | 0.796 [0.744, 0.84] | ref |
| | Yes | 7 (0:5; 1:2) | | | |
| Age | [0,25] | 33 (0:22; 1:11) | | | |
| | (25,35] | 123 (0:88; 1:35) | 1 [0.901, 1] | 0.034 [0.012, 0.095] | ref |
| | (35,45] | 133 (0:98; 1:35) | 0.943 [0.814, 0.984] | 0.031 [0.01, 0.086] | 0.94 [0.87, 1.02] |
| | (45,70] | 75 (0:57; 1:18) | 1 [0.824, 1] | 0.035 [0.01, 0.119] | 1 [0.9, 1.08] |

| | | | | | |
|---|---|---|---|---|---|
| Menopause | No | 319 (0:229; 1:90) | 0.967 [0.907, 0.989] | 0.026 [0.012, 0.056] | ref |

### S15. Supplementary material 15: Investigation of interaction between patient characteristics on the association between diagnostic test and disease status

#### A. Interaction between patient characteristics and HPV

| | **Crude** | | | | **Adjusted** | | | | |
|---|---|---|---|---|---|---|---|---|---|
| | **N** | **Odds Ratios** | **CI** | **Pval** | **N** | **Odds Ratios** | **CI** | **Pval** | **Adj. model** |
| HPV x Age (per 10 years) | 363 | 1.363 | [0.783, 2.4] | 0.28 | | | | | |
| HPV x CD4 count [cells/mm3] (per 1000 cells) | 368 | 0.299 | [0.047, 1.976] | 0.2 | | | | | |
| HPV x Contraception use | 369 | 1.036 | [0.379, 2.884] | 0.95 | | | | | |
| HPV x Education | 369 | 0.759 | [0.208, 3.029] | 0.68 | | | | | |
| HPV x History of treatment for precancer | | | | | | | | | |
| HPV x HIV RNA load [copies/mL] | 369 | 2.761 | [0.37, 58.118] | 0.39 | | | | | |
| HPV x Menopause | 369 | 0.635 | [0.148, 2.829] | 0.54 | | | | | |
| HPV x Parity | 369 | 0.842 | [0.656, 1.08] | 0.18 | | | | | |
| HPV x Trichomoniasis result | 369 | 0.972 | [0.297, 3.376] | 0.96 | | | | | |

B. Interaction between patient characteristics and VIA

| | Crude | | | | Adjusted | | | | |
|---|---|---|---|---|---|---|---|---|---|
| | **N** | **Odds Ratios** | **CI** | **Pval** | **N** | **Odds Ratios** | **CI** | **Pval** | **Adj. model** |
| VIA x Age (per 10 years) | 364 | 1.642 | [0.744, 3.728] | 0.22 | | | | | |
| VIA x CD4 count [cells/mm3] (per 1000 cells) | 370 | 0.376 | [0.03, 3.991] | 0.43 | | | | | |
| VIA x Contraception use | 371 | 0.950 | [0.205, 4.395] | 0.95 | | | | | |
| VIA x Education | 371 | 4.144 | [0.671, 35.443] | 0.14 | 363 | 3.941 | [0.629, 34.062] | 0.16 | out1 ~ education_level.factor3 + via + parity_nr + education_level.factor3:via |
| VIA x History of treatment for precancer | | | | | | | | | |
| VIA x HIV RNA load [copies/mL] | 371 | 0.619 | [0.096, 4.367] | 0.62 | | | | | |
| VIA x Menopause | 371 | 0.514 | [0.021, 6.462] | 0.61 | | | | | |
| VIA x Parity | 371 | 1.009 | [0.715, 1.409] | 0.96 | | | | | |
| VIA x Trichomoniasis result | 371 | 1.969 | [0.331, 16.305] | 0.48 | 363 | 4.767 | [0.6, 101.918] | 0.19 | out1 ~ STI_BL + via + parity_nr + hiv_rna_cat_subgrp + STI_BL:via |

C. Interaction between patient characteristics and Gynocular (using Youden cut-off)

| | Crude | | | | Adjusted | | | | |
|---|---|---|---|---|---|---|---|---|---|
| | **N** | **Odds Ratios** | **CI** | **Pval** | **N** | **Odds Ratios** | **CI** | **Pval** | **Adj. model** |
| x Age (per 10 years) | 364 | 1.224 | [0.701, 2.145] | 0.48 | | | | | |
| x CD4 count [cells/mm3] (per 1000 cells) | 370 | 1.508 | [0.213, 11.086] | 0.68 | | | | | |
| x Contraception use | 371 | 1.729 | [0.618, 4.911] | 0.3 | | | | | |
| x Education | 371 | 0.390 | [0.101, 1.465] | 0.16 | | | | | |
| x History of treatment for precancer | | | | | | | | | |
| x HIV RNA load [copies/mL] | 371 | 0.393 | [0.08, 2.121] | 0.26 | | | | | |
| x Menopause | 371 | 0.791 | [0.171, 3.656] | 0.76 | | | | | |
| x Parity | 371 | 1.212 | [0.942, 1.566] | 0.14 | 363 | 1.237 | [0.961, 1.603] | 0.1 | out1 ~ parity_nr + swede_cat1 + parity_nr:swede_cat1 |
| x Trichomoniasis result | 371 | 0.915 | [0.276, 3.14] | 0.89 | | | | | |
